# Differences in polygenic score distributions in European ancestry populations: implications for breast cancer risk prediction

**DOI:** 10.1101/2024.02.12.24302043

**Authors:** Kristia Yiangou, Nasim Mavaddat, Joe Dennis, Maria Zanti, Qin Wang, Manjeet K. Bolla, Mustapha Abubakar, Thomas U. Ahearn, Irene L. Andrulis, Hoda Anton-Culver, Natalia N. Antonenkova, Volker Arndt, Kristan J. Aronson, Annelie Augustinsson, Adinda Baten, Sabine Behrens, Marina Bermisheva, Amy Berrington de Gonzalez, Katarzyna Białkowska, Nicholas Boddicker, Clara Bodelon, Natalia V. Bogdanova, Stig E. Bojesen, Kristen D. Brantley, Hiltrud Brauch, Hermann Brenner, Nicola J. Camp, Federico Canzian, Jose E. Castelao, Melissa H. Cessna, Jenny Chang-Claude, Georgia Chenevix-Trench, Wendy K. Chung, NBCS Collaborators, Sarah V. Colonna, Fergus J. Couch, Angela Cox, Simon S. Cross, Kamila Czene, Mary B. Daly, Peter Devilee, Thilo Dörk, Alison M. Dunning, Diana M. Eccles, A. Heather Eliassen, Christoph Engel, Mikael Eriksson, D. Gareth Evans, Peter A. Fasching, Olivia Fletcher, Henrik Flyger, Lin Fritschi, Manuela Gago-Dominguez, Aleksandra Gentry-Maharaj, Anna González-Neira, Pascal Guénel, Eric Hahnen, Christopher A. Haiman, Ute Hamann, Jaana M. Hartikainen, Vikki Ho, James Hodge, Antoinette Hollestelle, Ellen Honisch, Maartje J. Hooning, Reiner Hoppe, John L. Hopper, Sacha Howell, Anthony Howell, ABCTB Investigators, kConFab Investigators, Simona Jakovchevska, Anna Jakubowska, Helena Jernström, Nichola Johnson, Rudolf Kaaks, Elza K. Khusnutdinova, Cari M. Kitahara, Stella Koutros, Vessela N. Kristensen, James V. Lacey, Diether Lambrechts, Flavio Lejbkowicz, Annika Lindblom, Michael Lush, Arto Mannermaa, Dimitrios Mavroudis, Usha Menon, Rachel A. Murphy, Heli Nevanlinna, Nadia Obi, Kenneth Offit, Tjoung-Won Park-Simon, Alpa V. Patel, Cheng Peng, Paolo Peterlongo, Guillermo Pita, Dijana Plaseska-Karanfilska, Katri Pylkäs, Paolo Radice, Muhammad U. Rashid, Gad Rennert, Eleanor Roberts, Juan Rodriguez, Atocha Romero, Efraim H. Rosenberg, Emmanouil Saloustros, Dale P. Sandler, Elinor J. Sawyer, Rita K. Schmutzler, Christopher G. Scott, Xiao-Ou Shu, Melissa C. Southey, Jennifer Stone, Jack A. Taylor, Lauren R. Teras, Irma van de Beek, Walter Willett, Robert Winqvist, Wei Zheng, Celine M. Vachon, Marjanka K. Schmidt, Per Hall, Robert J. MacInnis, Roger L. Milne, Paul D.P. Pharoah, Jacques Simard, Antonis C. Antoniou, Douglas F. Easton, Kyriaki Michailidou

## Abstract

The 313-variant polygenic risk score (PRS_313_) provides a promising tool for breast cancer risk prediction. However, evaluation of the PRS_313_ across different European populations which could influence risk estimation has not been performed. Here, we explored the distribution of PRS_313_ across European populations using genotype data from 94,072 females without breast cancer, of European-ancestry from 21 countries participating in the Breast Cancer Association Consortium (BCAC) and 225,105 female participants from the UK Biobank. The mean PRS_313_ differed markedly across European countries, being highest in south-eastern Europe and lowest in north-western Europe. Using the overall European PRS_313_ distribution to categorise individuals leads to overestimation and underestimation of risk in some individuals from south-eastern and north-western countries, respectively. Adjustment for principal components explained most of the observed heterogeneity in mean PRS. Country-specific PRS distributions may be used to calibrate risk categories in individuals from different countries.

## Introduction

Genetic susceptibility to breast cancer is influenced by multiple genetic variants which contribute different levels of risk to the disease (1-6). Genome-wide Association Studies (GWAS) have identified thus far a large number of common, low-risk variants that each contribute a small risk to the disease but can be combined into Polygenic Risk Scores (PRSs) with larger effect (7, 8). PRSs provide a promising tool for clinical risk prediction of breast cancer by stratifying women into different categories of breast cancer risk (9-11), and may be used to inform targeted screening and prevention strategies (12-20).

Mavaddat et al., (2019) constructed a 313-variant PRS (PRS_313_) for breast cancer, using data for women of European ancestry from the Breast Cancer Association Consortium (BCAC) (11). In prospective validation studies, this PRS was estimated to be associated with a relative risk for breast cancer of approximately 1.6 per standard deviation increase, and its discriminatory ability, measured in terms of area under the ROC curve (AUC), was 0.63. The lifetime absolute risk of developing breast cancer for individuals in the lowest percentile of the PRS_313_ risk distribution was estimated to be ∼2%, while for those in the highest percentile it was ∼33%. PRS_313_ has been incorporated into the multifactorial BOADICEA (Breast and Ovarian Analysis of Disease Incidence and Carrier Estimation Algorithm) model which is available via the CanRisk tool (14, 21, 22) (www.canrisk.org) and, together with other lifestyle and genetic risk factors, has been shown to improve risk stratification in European and European ancestry populations (14, 23-27). PRS_313_ has also been shown to be transferrable to women of other ethnic backgrounds, although the strength of the association with breast cancer risk was attenuated compared with that for women of European ancestry (OR per SD (95% CI) 1.52 (1.49-1.56), AUC = 0.61 in women of east Asian ancestry; OR 1.27 (1.23-1.31), AUC = 0.57 in women of African ancestry (28-30)).

Although several studies have investigated the transferability of PRS developed in European ancestry populations to non-European populations (31-34), the PRS distributions across different European countries has not been extensively evaluated. Differences in the PRS distribution, if not appropriately accounted for, could lead to inappropriate risk classification, with implications for clinical management.

In this study, we aimed to examine the distribution of the PRS_313_ across 17 countries in Europe, together with individuals of European ancestry from Australia, Canada, Israel and the USA. Similar analyses were performed using data from the UK Biobank, stratifying individuals by country of birth. We explored different approaches to account for the differences in the distribution, and investigated the implications of distribution differences across countries in breast cancer risk prediction.

## Materials and methods

### Study populations and Genotyping

#### Breast Cancer Association Consortium dataset

The BCAC dataset used here consisted of 110,260 female invasive breast cancer cases and 94,072 female healthy controls of European ancestry, recruited into 84 studies from 21 countries participating in the BCAC (**Supplementary Table 1A**). For simplicity and with attempt to explore the effect on the general female population, only the control data were used in these analyses as the distribution of the PRS in cases might vary between studies due to differences in study design (in particular oversampling of cases with a family history of disease). Samples from participating individuals were genotyped using the iCOGS (1) or OncoArray (3, 35) genotyping array. For samples genotyped using both arrays, the OncoArray genotype data were used. The iCOGS and the OncoArray datasets were imputed separately in a two-step manner using SHAPEIT (36) for phasing and IMPUTE2 for imputation. The Phase 3 (October 2014) release of the 1000 Genomes data (37) was used as the reference panel. More details on genotyping, quality control and imputation are given elsewhere (2, 3, 35). Ancestry-informative principal components (PCs), derived separately from the iCOGS and the OncoArray genotypes, were also calculated for all the samples, as previously described (3).

**Table 1:** Mean standardized PRS_313_ by country in controls in the pooled BCAC dataset, estimated when adjusted for array, 6 PCs country and array, using fitted values adjusted for 6 PCs and array and when using an Empirical Bayes approach adjusted for array.

| Country | Number of Controls | Mean PRS<sub>313</sub><sup>1</sup> | Mean PRS adjusted for array and 6 PCs | PRS adjusted for 6 PCs, fitted values<sup>2</sup> | Empirical Bayes Posterior Mean<sup>3</sup> |
| --- | --- | --- | --- | --- | --- |
| Australia | 4049 | -0.005 | 0.01 | -0.005 | -0.003 |
| Belarus | 342 | 0.07 | 0.071 | 0.016 | 0.064 |
| Belgium | 1823 | -0.006 | -0.007 | 0.010 | -0.002 |
| Canada | 2277 | 0.018 | 0.019 | 0.013 | 0.02 |
| Denmark | 5241 | -0.013 | 0.012 | -0.031 | -0.012 |
| Finland | 2083 | 0.031 | 0.008 | 0.010 | 0.032 |
| France | 1372 | 0.0003 | -0.008 | 0.008 | 0.004 |
| Germany | 8563 | 0.011 | 0.004 | 0.013 | 0.011 |
| Greece | 607 | 0.232 | 0.043 | 0.208 | 0.199 |
| Ireland | 719 | -0.118 | -0.015 | -0.112 | -0.092 |
| Israel | 724 | 0.047 | 0.001 | 0.062 | 0.047 |
| Italy | 1554 | 0.115 | -0.007 | 0.131 | 0.11 |
| Netherlands | 4407 | 0.021 | 0.043 | -0.019 | 0.022 |
| Norway | 217 | 0.077 | 0.094 | -0.027 | 0.066 |
| Poland | 2554 | 0.013 | 0.025 | 0.010 | 0.015 |
| Republic of North Macedonia | 92 | 0.25 | 0.134 | 0.140 | 0.129 |
| Russia | 120 | 0.18 | 0.166 | 0.044 | 0.11 |
| Spain | 2098 | 0.057 | -0.006 | 0.057 | 0.056 |
| Sweden | 16680 | -0.015 | 0.005 | -0.017 | -0.014 |
| UK | 16854 | -0.01 | 0.019 | -0.023 | -0.01 |
| USA | 21696 | 0.029 | 0.033 | 0.013 | 0.029 |

<sup>1</sup>Mean PRS<sub>313</sub> adjusted for array
<sup>2</sup>Mean PRS<sub>313</sub> by country using predicted PRS of each individual; estimated using linear predictor of PRS vs 6 PCs and the command predict () in R.
<sup>3</sup>Country-specific estimates, means $\beta$ , using the Empirical Bayes approach, adjusted for array

#### UK Biobank dataset

UK Biobank, is a prospective cohort study including more than 500,000 participants from England, Wales and Scotland, with age at recruitment between 40 to 69 years old, more details can be found elsewhere (38, 39). For the analyses in this study, genotype data from females (genetic reported sex) participating in the UK Biobank were used. Individuals were excluded if they had a recorded breast cancer diagnosis (malignant neoplasm of breast or carcinoma in situ of breast) or had a personal history of malignant neoplasm of breast. Genetic ancestry was inferred using the FastPop software (40). Individuals self-reported “white” and with an estimated European ancestry proportion ≥ 80% were retained in the analysis. Then, individuals were stratified by the “country of birth” field in the UK Biobank; only countries with at least 100 participants were used. After filtering, 225,105 females from 17 countries of Europe and from Australia, Canada, New Zealand, and the USA were used in the analyses (more details in **Supplementary Table 1B)**. Samples were genotyped using the Affymetrix UK BiLEVE Axiom array and the Affymetrix UK Biobank Axiom array. Imputation data used were based on the Haplotype Reference Consortium (41), the UK10K +1000 Genomes panel references. More details on genotyping, quality control and imputation are given elsewhere (38). Ancestry informative PCs were also available (38).

All study participants gave written informed consent, and all the studies were approved by the relevant ethics committees. The use of the UK Biobank has been approved under application ID102655.

### Statistical Analyses

PRS_313_ was developed previously using a hard-thresholding stepwise forward regression approach, and included variants independently associated with breast cancer risk at a p-value cut off < 10^−5^ (11). PRS_313_ was calculated in each study participant using the following formula:

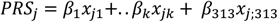

Where *PRS*_*j*_ is the PRS of individual j, *x*_*jk*_ is the estimated effect allele dosage for *SNP*_*k*_ carried by individual *j* and can take values between 0 and 2, and *β*_*k*_ is the weight for *SNP*_*k*_ in the PRS for overall breast cancer, as derived by Mavaddat et al. (11). PRS_313_ was standardized to have unit SD in controls in the pooled dataset. Mavaddat et al. (11) also derived ER-specific versions of PRS_313_, with weights optimised for predicting ER-positive or ER-negative breast cancer risk (**Supplementary Table 2**).

**Table 2:**
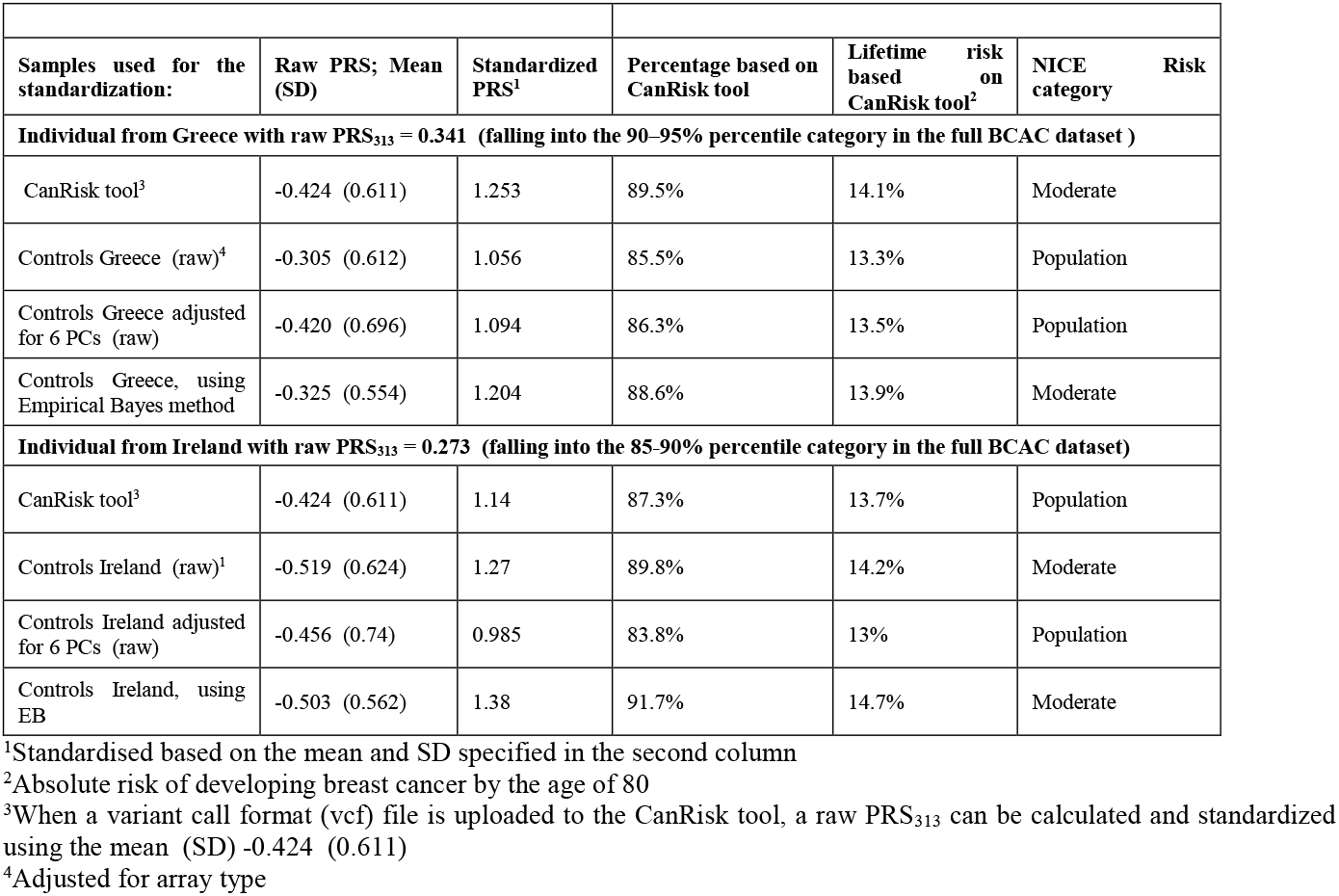
Mean and SD used to standardize PRS_313_ of a 50-year-old woman with raw PRS_313_ equal to 0.341 from Greece and another 50-year-old woman with raw PRS_313_ equal 0.273 from Ireland, and the risk estimation and categorization when using the CanRisk tool, Greek and Ireland values

The main analyses focused on calculating the mean standardized PRS_313_ in BCAC controls, using both the iCOGS and OncoArray datasets. These values were derived using linear regression with array type as a covariate and no intercept (so that estimates were generated for every country). Heterogeneity in the mean PRS_313_ between countries was assessed using I^2^ statistics and Q statistic p-values.

We also evaluated the distribution of the mean PRS by country of birth in female participants in the UK Biobank. Seven of the 313 variants were not available from the UK Biobank data and thus we used the remaining 306 variants in the analysis (PRS_306_) (**Supplementary Table 2**). We also evaluated a “standard” breast cancer PRS available in the UK Biobank data, previously generated from external GWAS data (42), and was available for 224,776 individuals (**Supplementary Table 1B**).

Potential sources of the variability in the mean PRS_313_ across the countries were explored in the BCAC dataset using three approaches. The PRS was first recalculated excluding variants in the *CHEK2* region. The protein truncating variant *CHEK2* c.1100delC is a relatively common founder variant that exhibits a large variation in frequency across Europe (43). Although it is not included in PRS_313_, other variants in the PRS_313_ are correlated with this variant. For this reason, the four variants in the *CHEK2* region included in PRS_313_ (the *CHEK2* p. Ile157Thr variant, and variants at positions 29135543, 29203724 and 29551872 on chromosome 22, positions based on build 37) were removed, resulting in a 309-variant PRS (PRS_309_). Mean and SE by country were recalculated for PRS_309_, as described above.

Second, we examined the effect of removing variants with the most variable frequency across countries. For this analysis, the mean and SD of the effect allele frequency in controls of the pooled dataset was calculated for each of the 313 variants by country. Variants with a coefficient of variation (SD/mean) greater than 0.3 were removed. Means and SE of the newly constructed PRS were recalculated by country as described above.

Third, we explored the effect of adjusting for up to 10 ancestry-informative PCs, in addition to type of array. As the PCs derived from the iCOGS and OncoArray are not comparable, separate PCs for each were included in the regression. We explored the number of PCs that were required to eliminate the heterogeneity in the adjusted mean PRS_313_, using the thresholds I^2^ < 10% and p-value > 0.05. Similarly, for the UK Biobank dataset, PRS_306_ was adjusted for up to 10 PCs, which were available in the UK Biobank.

As a complementary approach to generating population-specific estimates, we explored an empirical Bayes approach similar to that described by Clayton and Kaldor (44) for mapping disease rates. The motivation of this approach is that, if some of the variation in means among countries is genuine, while some is due to sampling variation, better estimates of the country-specific means can be obtained by “shrinking” the country-specific estimates towards the overall mean, by an amount depending on the sample size. In our implementation, we allowed the PRS means to be correlated between countries, using the autocorrelation matrix proposed in Clayton and Kaldor. A detailed description is given in **Supplementary Methods**.

To investigate the implications of PRS distribution differences in breast cancer risk prediction, we explored the proportion of women by country by percentile (<1%, 1%-5%, 5%-10%, 10%-20%, 20%-40%, 40%-60%, 60%-80%, 80%-90%, 90%-95%, 95%-99%, ≥99% percentiles), based on the distribution cut-offs of either the full dataset or country-specific estimates. We also examined a specific risk estimation example using the CanRisk tool (14, 21, 22).

All analyses were performed in R (version 4.2.1) (45). Forest plots were generated using the metafor package (46). Maps were generated using the packages world map data from natural earth (rnaturalearth) (47), the world vector map data from natural earth used in ‘rnaturalearth’ (rnaturalearthdata) (48), simple features for R (sf) (49) and interface to geometry engine (rgeos) (50).

## Results

### Geographic diversity in the mean PRS_**313**_ **across European ancestry populations**

The mean PRS_313_ in the BCAC controls differed markedly across European countries, with heterogeneity I^2^ = 80% (p-value = 5.6 × 10^−13^). The mean was highest in the Republic of North Macedonia (0.25), Greece (0.23), Russia (0.18) and Italy (0.12), and lowest in Ireland (−0.12). The mean estimates for Australia, Canada, Israel and the USA were close to the overall mean (**Figure 1; Figure 2; Table 1; Supplementary Table 3A**). A similar level of heterogeneity was observed for the ER-positive (I^2^ = 84%) and ER-negative PRS (I^2^ = 64%) (**Figure 2; Supplementary Table 3B**). There was no evidence of a difference in the SD of the PRS between countries. (**Supplementary Table 3A**).

**Figure 1:**
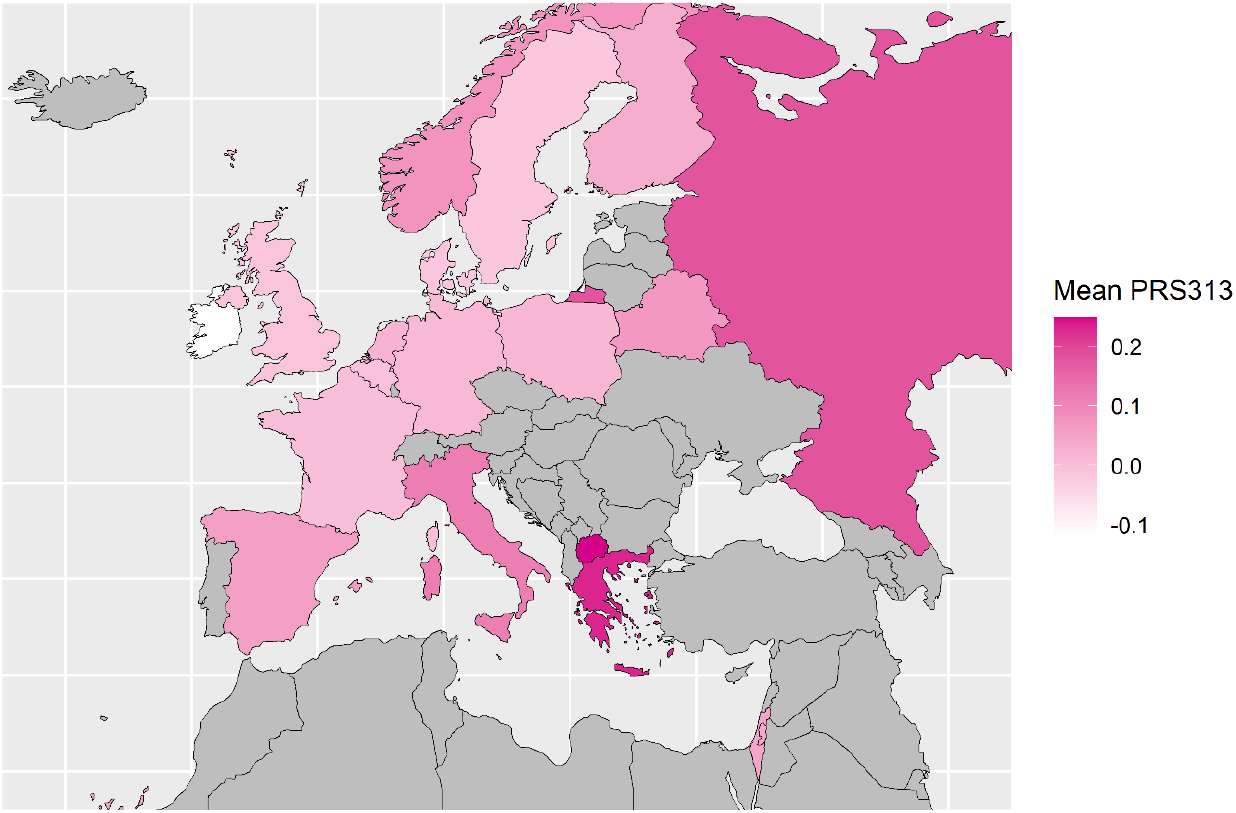
Map of the European countries of origin of BCAC study participants included in the analysis. Countries were coloured based on their mean standardized PRS_313_ in control dataset of BCAC. Countries with higher mean are represented with darker colour while those with lower mean with lighter colour.

**Figure 2:**
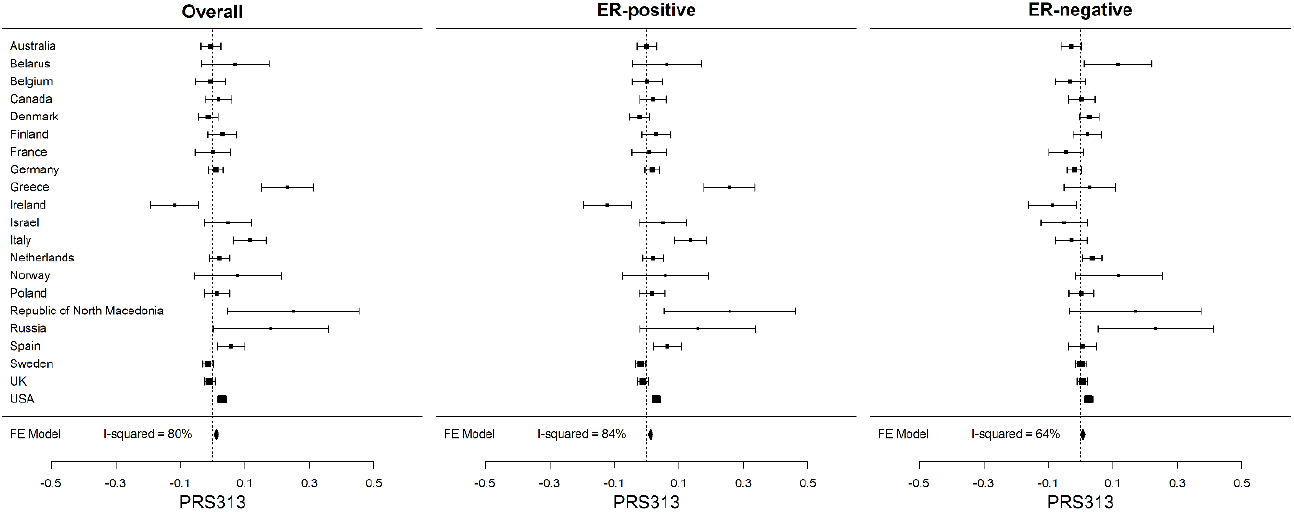
Distribution of the standardized PRS_313_ across country of origin for overall, ER-positive and ER-negative breast cancer in control dataset of BCAC. The squares represent the mean PRS by country and the error bars represent the corresponding 95% confidence intervals (FE Model: Fixed effect Model).

The mean PRS_306_ in female UK Biobank participants, stratified by country of birth, was also calculated (**Figure 3** and **Supplementary Table 4)**. There was strong evidence of heterogeneity in the PRS distribution (I^2^ = 66%, p-value = 2.3 × 10^−06^). The pattern was generally similar to that seen in the BCAC dataset, with a higher PRS in individuals born in southern and eastern Europe (e.g. Cyprus, Russia, Italy) and lower in western Europe (e.g. Ireland). Similar results were found for the “standard” UK Biobank PRS (I^2^ = 87%, p-value = 1.4 × 10^−25^) (**Figure 3 and Supplementary table 4**).

**Figure 3:**
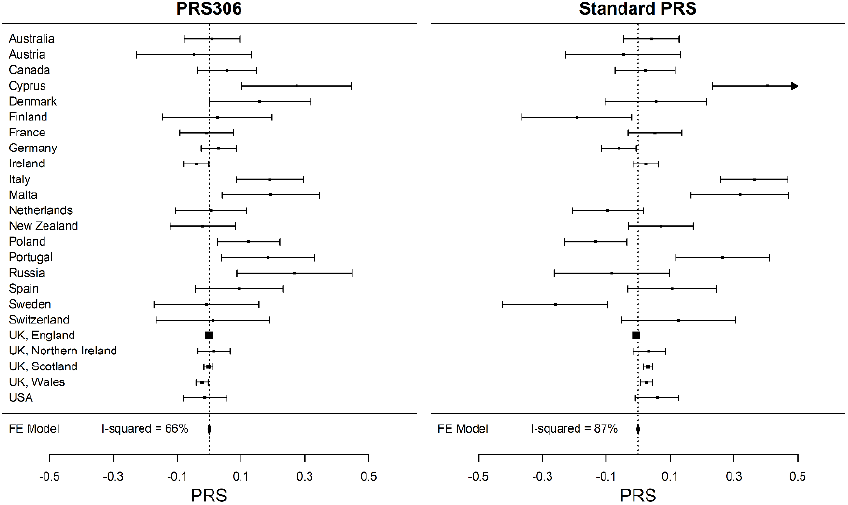
Distribution of the mean PRS_306_, and “standard” PRS for breast cancer, as defined in the UK Biobank, across countries of origin of participating white females. The squares represent the mean PRS by country and the error bars represent the corresponding 95% confidence intervals (FE Model: Fixed effect Model).

### Exploring potential reasons for differences in mean standardized PRS between countries

Potential sources of the variability in the mean PRS_313_ across the countries were explored in the BCAC dataset, using three approaches. We first evaluated the effect of removing variants in the *CHEK2* region on the distribution of the mean PRS_313_ for the countries. After removing these four variants, the variation in the mean PRS_309_ across countries in the controls remained similar that for PRS_313_ (I^2^ = 83%, p-value = 9.4 × 10^−16^). We next identified the variants with the most variable frequency from countries in the control dataset. Seventeen of the 313 variants had a coefficient of variation greater than 0.3 (**Supplementary Table 5**). Excluding these 17 variants did not reduced the variation in the mean PRS (I^2^ = 80%, p-value = 2.4 × 10^−12^).

We next explored the effect of adjusting for PCs. When individuals in the BCAC dataset genotyped with OncoArray were plotted by the first two PCs, those from the same country separated clearly, in a pattern consistent with their geographical relationship (**Supplementary Figure 1**). This suggests that adjusting for PCs maybe an effective approach to reducing the variation in PRS distribution. When we adjusted the PRS for the leading PCs in the BCAC dataset, the I^2^ reduced as each PC was added in the model and reached < 10% when adjusted for the first six PCs (I^2^ = 69%, 54%, 47%, 39%, 22%, 0%, and 0% when including 1, 2, 3, 4, 5, 6, and 10 PCs respectively) (**Table 1**; **Supplementary Table 3A; Supplementary Figure 2**). A similar result was obtained for the ER-positive PRS (**Supplementary Table 3B**), when adjusted for the first 6 PCs (Heterogeneity: I^2^ = 0%, p-value = 0.69). For the ER-negative PRS, however, the heterogeneity was not eliminated even when the PRS was adjusted for 10 PCs (Heterogeneity: I^2^ = 56%, p-value = 0.001) (**Supplementary Table 3B**). The predicted PRS of each individual, as derived from the fitted values of the linear regression model of PRS adjusted for the first 6 PCs and array, were then used to calculate a predicted mean PRS_313_ by country (**Table 1** and **Supplementary Table 3A**).

We repeated these analyses for PRS_306_ using the UK Biobank dataset. I^2^ reduced as each PC was added in the model and reached < 10% when adjusted for the first eight PCs (**Supplementary Table 4** and **Supplementary Figure 3**).

### Mean PRS estimates by country calculated using an Empirical Bayes approach

The empirical Bayes estimates by country for the mean PRS in controls of the BCAC dataset are given in **Table 1** and **Supplementary Table 6**. Compared with the unadjusted estimates, the estimates shrunk towards the overall mean, with the shrinkage being greatest for countries that had small available sample sizes, such as Republic of North Macedonia and Russia (**Table 1**). The adjusted mean PRS by country were generally similar to those predicted by the model adjusting for six PCs (**Supplementary Table 6**). When PRSs were adjusted for the first 6 PCs, applying the empirical Bayes approach makes little difference to the estimates (**Supplementary Table 6**).

### Implications for Breast Cancer Risk Prediction

To explore the effect of these differences in PRS distribution between different European populations on risk stratification, we first defined risks thresholds based on the distribution of the controls in the full BCAC dataset (**Supplementary Table 7A)**. We then calculated the percentage of controls by country that would be categorized in the 90-95^th^, 95-99^th^, and >99^th^ percentile categories, based on the distribution in the full dataset, and compared these to the percentages based on the country-specific distributions (**Supplementary Tables 7B-D**). Based on the overall distribution, approximately 4.1%, 3.7%, 1.3% and 0.5% additional women from Belarus, Republic of North Macedonia, Greece, and Italy, respectively, would be incorrectly classified in the 95-99^th^ percentile instead at the 90-95^th^ percentile; while 1.1% and 1.4% additional women from France and Ireland, respectively, would be incorrectly classified in the 90-95^th^ instead of the 95-99^th^ percentile (**Supplementary Table 7C**). **Figure 4** and **Supplementary Table 8** illustrate the PRS_313_ percentile distribution in the full dataset, Greece, Italy (countries with the highest PRS_313_ and including more than 100 controls) and Ireland (lowest PRS_313_).

**Figure 4:**
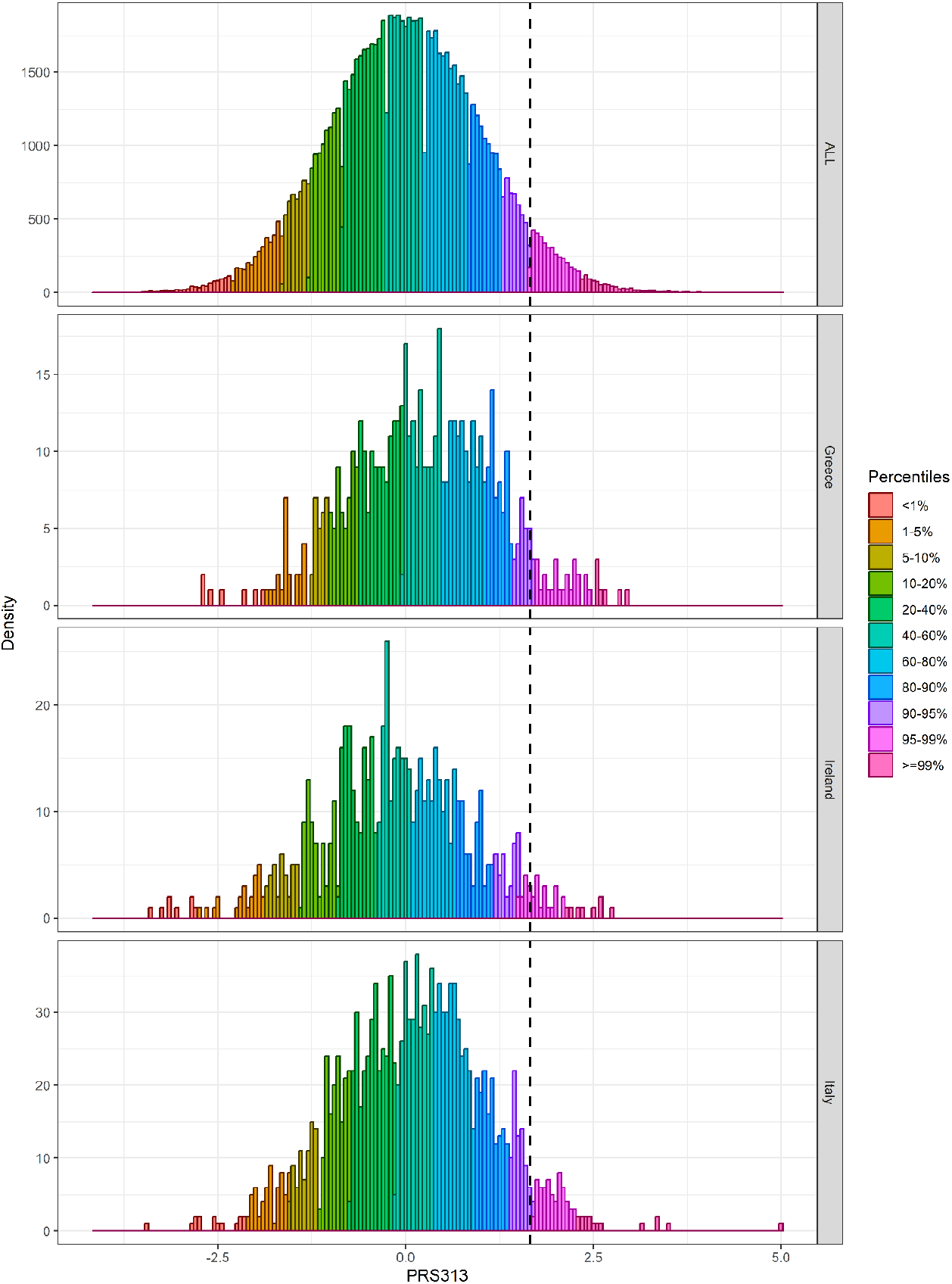
PRS_313_ distribution in controls by percentiles in the pooled BCAC dataset, Greece, Ireland and Italy. The dashed line corresponds to the 95th percentile of the PRS_313_ distribution in controls of the pooled BCAC dataset.

We next considered as an example a 50-year-old female from Greece with a raw PRS_313_ of 0.3414 (falling into the 90^th^ – 95^th^ percentile category in the full BCAC dataset) and no data on family history or other known risk factors. As Greek incidence rates are not available and not currently implemented in CanRisk, we used the UK incidence rates for the calculations. If this PRS was standardized based on the mean and SD used in the CanRisk tool (when a variant call format (vcf) file is uploaded to the CanRisk tool, a raw PRS_313_ can be calculated and standardized using the mean: -0.424; SD: 0.611), the individual would (assuming UK incidence rates) be given an estimate of 14.1% risk of developing breast cancer by the age of 80 and classified in the moderate risk category (**Table 2**). On the other hand, if the PRS were standardized based on the mean and SD of the controls of Greece (mean: -0.305; SD: 0.612-raw values), she would fall in the 80-90% percentile category with an estimated 13.3% risk of developing breast cancer by the age of 80, and be classified into the population risk category (**Figure 5** and **Table 2**). Similarly, if the PRS were standardized based on the mean and SD of PRS for Greece predicted by adjustment for the first 6 PCs (mean: -0.42, SD: 0.696), she would also be classified in the population risk category (**Table 2**). Finally, if the PRS were standardized based on the mean and SD of the empirical Bayes approach (mean: -0.325 SD: 0.554), will have an estimated 13.9% risk of developing breast cancer by the age of 80, and be classified into the moderate risk category (**Table 2**).

**Figure 5:**
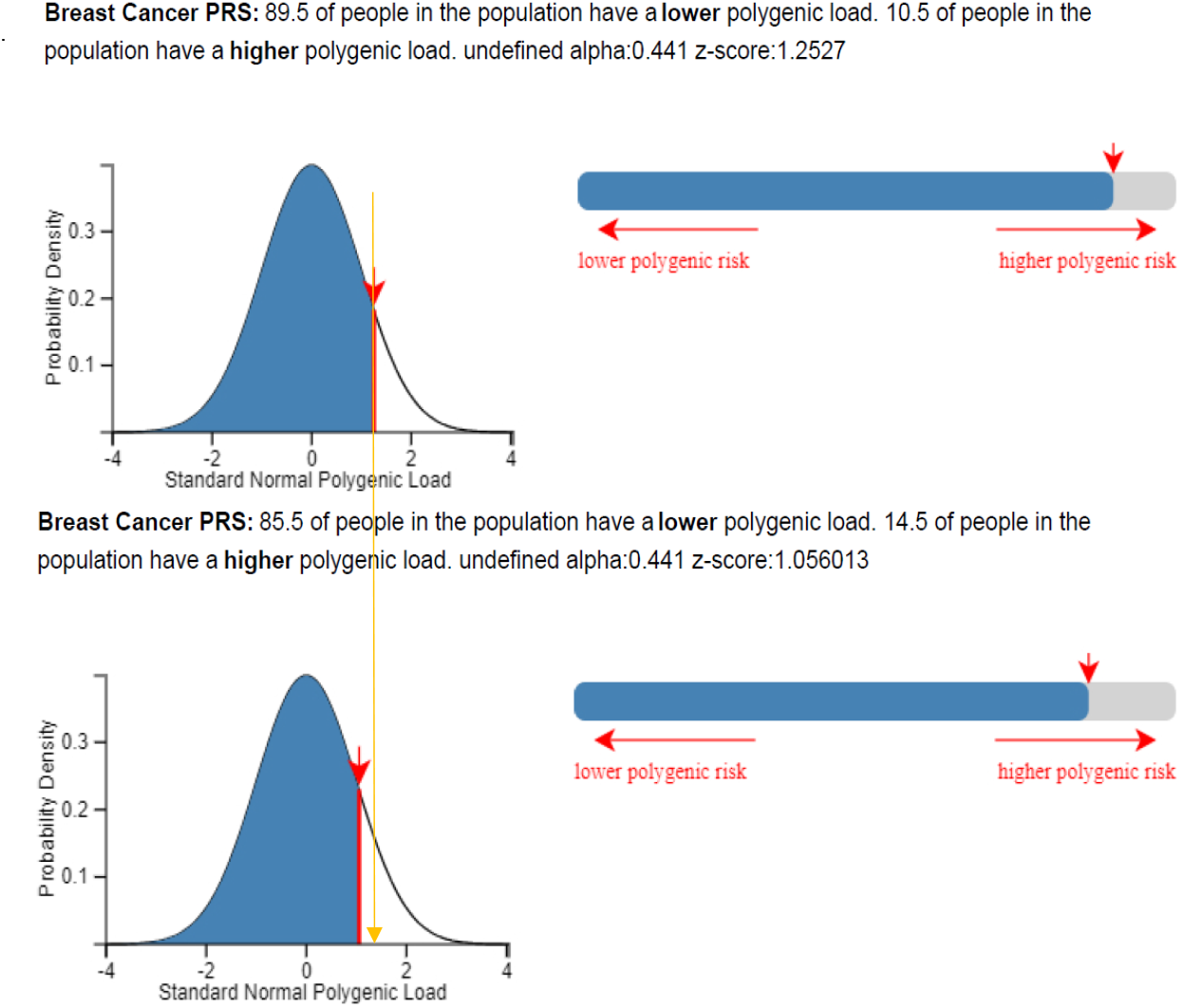
Classification of a 50-year-old woman from Greece when her raw PRS_313_, which is equal to 0.34 is standardized based on the mean and SD of the controls of BOADICEA model (upper panel) and Greece (lower panel), using the CanRisk tool. Plots were generated using the CanRisk tool (www.canrisk.org).

A second example is illustrated in Table 2 and Figure 6, based on a 50-year-old female from Ireland with raw PRS_313_ equal to 0.273 (equivalent to the 85^th^ – 90^th^ percentile-in the full BCAC dataset) and no other risk factors known. Using the CanRisk tool and assuming UK incidence rates, she would be classified in the 87.3% percentile with an estimated 13.7% absolute risk of developing breast cancer by the age of 80, which according to the NICE guidelines would be classified in the population risk category. If the PRS was standardized based on the mean and SD of PRS_313_ as derived from the controls in Ireland (mean for Ireland: -0.519, and SD: 0.624 -raw values), then she would be classified in the 89.8% percentile with estimated 14.2% risk of developing breast cancer by the age of 80, classified in the moderate risk category (**Figure 6**). If the PRS were standardized based on the mean and SD of PRS for Ireland predicted by adjustment for the first 6 PCs (mean: -0.456, SD: 0.74), she would also be classified in the population risk category (**Table 2**). Finally, if the PRS were standardized based on the mean and SD of the empirical Bayes approach (mean: -0.503, SD: 0.562), will have an estimated 14.7% risk of developing breast cancer by the age of 80, and be classified into the moderate risk category (**Table 2**).

**Figure 6:**
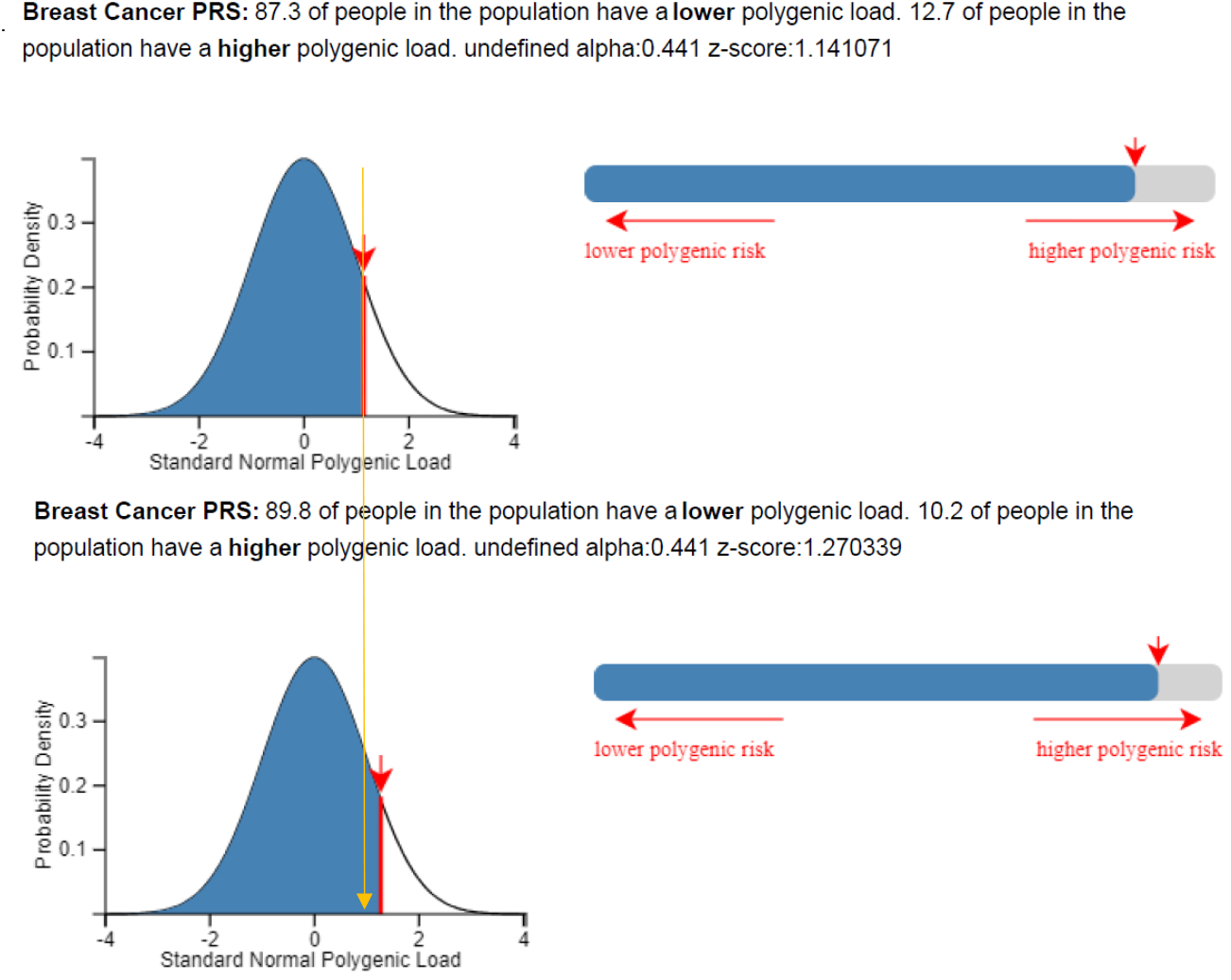
Classification of a 50-year-old woman from Ireland when her raw PRS_313_, which is equal to 0.27 is standardized based on the mean and SD of the controls BOADICEA (upper panel) model and Ireland (lower panel), using the CanRisk tool. Plots were generated using the CanRisk tool (www.canrisk.org).

## Discussion

Transferability of PRSs across different populations remains a major challenge in the field of personalized cancer risk prediction (31, 51). In this study, we explored the distribution of PRS_313_ for breast cancer in European ancestry women from 21 countries, using data from studies participating in the BCAC, and further investigated how the observed variability might be accounted for in breast cancer risk prediction.

The results indicated that the PRS_313_ distribution varies markedly even within Europe, with a higher mean in south-east Europe (e.g. Republic of North Macedonia, Greece, Italy) and a lower mean in western Europe (e.g. Ireland). We observed a very similar pattern in females participating in the UK Biobank, based on country of birth. If not accounted for, these differences would lead to an over- or under-estimation of risk, thus affecting the risk categorization and possibly the clinical management of some women. This may be important not only at the individual country level but also for individuals living in a different country to that of their origin.

The variability in the mean PRS_313_ could not be explained by removing variants with the most variable frequency, indicating that a large number of variants may contribute to this difference. Removing such variants to reduce the heterogeneity would not in any case be desirable as it would reduce the risk discrimination provided by the PRS. The results do, however, indicate that most if not all of the variability in the mean PRS_313_ across countries in controls can be explained by adjusting for the leading ancestry informative PCs (6 PCs in the BCAC datasets, based on the OncoArray or iCOGS arrays, 8 PCs in UK Biobank). An advantage of using PCs is that they do not require any prior data from the population in question. A disadvantage, however, is that PCs require array genotyping data to generate, making them less attractive when implemented using sequencing panels. Moreover, the PCs generated using different genotyping arrays are not necessarily comparable. One interesting observation is that the heterogeneity of the ER-negative specific PRS was not eliminated even with the adjustment for 10 PCs.

We also explored generating country-specific mean PRS using an empirical Bayes approach. This approach considers both the uncertainty due to the small available sample size and the true variation in the means across the countries; these country-specific mean PRS were similar to those generated by adjusting for PCs. These values can then be used to standardise the PRS before, for example, implementing in the CanRisk tool. The risk categorization of the females from Greece and Ireland, the two examples in the Result section, was changed depending on the mean and SD of the sample used for the standardization of PRS. According to the NICE, women classified in the moderate risk category (lifetime risk of at least 17% and less than 30%), have different managing guidelines compared to women classified in the population risk category (52).

While adjustment of the PRS distribution at the population level is clearly necessary, the results raise the question as to whether it is appropriate in general to adjust PRS for PCs at the individual level, which gives different scores and potentially different risk classifications. This is a difficult question to address and hinges on whether the PCs should be regarded as nuisance parameters correcting for confounding factors, such as screening or lifestyle factors. Reanalysis of prospective studies with BCAC OncoArray dataset shows that the first two PCs are associated with the PRS (PC1 negatively, PC2 positively) and are also associated with risk (in the same direction). The PRS effect size (OR per 1 SD) is essentially unchanged whether or not adjustment for PCs is made (Supplementary Table 9 and Supplementary Table 10). This implies that risk discrimination would be slightly improved by including the effect of PCs in the PRS, and that adjusting the PRS for PCs further reduces the discrimination. Fortunately, the association between the PC1 and risk is weak and, within a country, the variation in the PC1 is not large enough to materially change risk categories.

The differences in the PRS distribution across Europe are a manifestation, on a continental scale, of the larger intercontinental differences – the mean PRS is higher in both east Asian and African populations than in the European dataset examined here (28, 29, 53). It is interesting to note that the pattern appears unrelated to the population-specific incidence, which is fact lower in south-east than north-west Europe (54), presumably because the effect on disease incidence is counterbalanced by larger effects of lifestyle (or other genetic) factors. It remains unclear whether the differences in the PRS can be attributed purely to random genetic drift or whether selection pressures relevant to breast cancer aetiology are involved.

We would like to acknowledge some potential limitations of our study. The dataset we used was genetically-homogeneous and maybe not completely representative of the population of each country. It remains an important issue how to interpret the PRS in individuals classified as mixed ancestry. In the future, the exploration of the distribution of the mean PRS across the individuals classified as mixed ancestry could be performed. Furthermore, evaluation of the country-specific calibrated PRS in combination with classical breast cancer risk factors should be performed in order to explore the extend to these findings have on final risk prediction.

In summary, these results demonstrate that the implementation of the PRS313 in risk prediction models such as CanRisk/BOADICEA could potentially require country-specific calibration. This can be achieved by genotyping a large control group to obtain population-specific means, by using a principal components adjustment, or the empirical Bayes approach described here.

## Supporting information

Acknowledgments

## Data Availability

Data from UK Biobank are available through application to UK Biobank. Data from the Breast Cancer Association Consortium (BCAC) used in the present study are available through application to the BCAC Data Access Co-ordinating Committee.

## Author contributions

Writing Group: K.Y., K.Mi., D.F.E., A.C.A., N.Ma, and J. Si.; Study design: K.Mi., D.F.E., A.C.A., N.Ma., J.Si., and K.Y.; Data management: M.K.B., Q.W.; Statistical Analysis: K.Y., N.Ma, J.D., M.Z., D.F.E., K.Mi.; Provided data: M.A., T.U.A., I.L.A., H.A-C., N.N.A., V.A., K.J.A., A.Au., A.Bat., S.Be., M.Berm., A.Ber., K.Bia., N.B., C.Bo., N.V.B., S.E.B., K.Br., H.Bra., H.Bre., N.J.C., F.C., J.E.C., J.C-C., G.C-T., W.K.C., NBCS Collaborators, S.V.C., F.J.C., A.Cox., S.S.C., K.Cz., M.B.D., P.D., T.D., A.M.D., D.M.E., A.H.E., C.En., M.E., DG.E., P.A.F., O.F., H.F., M.G-D., A.G-M., A.G-N., P.Gu., E.Hah., C.A.H., P.Hall., U.H., J.M.H., V.H., J.H., A.Hol., E.Hon., M.J.H., R.H., J.L.Ho., S.H., A.How., ABCTB Investigators, kConFab Investigators, S.J., A.Jak., H.J., N.J., R.Ka., E.K.K., C.M.Ki., S.Kou., V.N.K., J.V.L., D.La., F.Lej., A.Lin., M.Lus., R.J.M., A.Man., D.M., U.M., R.L.M., R.A.M., H.Ne., N.Ob., K.Of., T-W.P-S., A.V.P., C.P., P.Pe., P.D.P.P., G.Pi., D.P.K., K.Py., P.Ra., M.U.R., G.R., E.R., J.R, A.Ro., E.H.R., E.S., D.P.S., E.J.S., M.K.S., R.K.S., C.Sc., X-O.S., M.C.S., J.St., J.A.T., L.R.T., C.M.V, I.VDB., W.W., R.Wi., W.Z., J.Si., A.C.A, D.F.E.. All authors read and approved the final version of the manuscript.

