## Supplementary Methods and Figures for "Differences in polygenic score distributions in European ancestry populations: implications for breast cancer risk prediction"

##### Empirical Bayes approach

Use of polygenic risk scores (PRS) for risk prediction requires that they are correctly calibrated. However, the mean PRS can vary markedly by population, so this may require population-specific means to calibrate correctly. This is a classic mapping problem, where one wishes to take account of the true variation in the distribution while also allowing for uncertainty due to small sample sizes.

The basic idea is that the PRS variation by country is modelled as a random effect $\beta \sim N(\mu, \sigma^2)$. In the general case, these effects can be correlated, so than $\beta \sim N(\underline{\mu}, \Sigma)$. The estimates of the country-specific estimates are the posterior mean $\beta$, given the data. This has the effect of shrinking the population-specific means towards the overall estimates in an efficient manner, with the shrinkage being greatest for countries with a small sample size and least for the country with a large sample size.

The general approach follows that used by Clayton and Kaldor (1987) (1) for mapping disease rates, building on the approach proposed by James and Stein (1961) for estimating multiple normal means simultaneously. Schmidt et al (2016) (2) used a similar approach for mapping the variation in *CHEK2**1100delC frequencies. Here the outcome, PRS, is considered normally distributed, simplifying the problem. In these notes we generally follow the notation used by Clayton et al Kaldor (1987) with minor variations.

In addition, we wish to determine whether the variation in PRS means can be explained by fitting ancestry informative principal components: this is plausible since the variation is driven by variation in allele frequencies and is more marked between than within continental ancestries.

Thus, the general model that we wish to fit is of the form:

$$y_j = \phi^T z_j + \beta_{g(j)} + \varepsilon_j$$

Here $y_j$ is the PRS for individual j, $\beta_{g(j)}$ is the PRS mean for individual $j$, in country *g(j)*, $z_j$ are the additional covariate values for individual $j$, $\phi$ are the corresponding parameters and $\varepsilon_j$ are the residual errors (assumed independent and distributed $\varepsilon_j \sim N(\mu, \tau^2)$. Note that, since the PRS means $\beta_k$ are being estimated for all n countries, no overall mean parameter should be included in the model or there would be redundancy. The covariates $z$ can include principal components, but also case-control status (if data on both cases and controls are available), array etc. (Note that in Clayton and Kaldor $z$ and $\phi$ refer to country-covariates such that $\mu = z\phi$. Here $z$ and $\phi$ refer to individual covariates and $\mu$ are the country-specific means (given covariates $z = 0$).

A standard approach to estimating the parameters is via an EM algorithm, with the population-specific means $\beta$ being considered as missing data. This is based on the general function:

$$Q(\mu, \Sigma, \phi) = E_{\mu', \Sigma', \phi'}(l(\mu, \Sigma, \phi)|y)$$

Where $l(\mu, \Sigma, \phi)$ is the complete data log-likelihood, and $\mu', \Sigma', \phi'$ refer to the current parameter estimates. The algorithm proceeds via a iterative repetition of an E-step and an M-step

E-step – compute $Q(\mu, \Sigma, \phi)$ given the current parameter estimates

M-step – maximise $Q(\mu, \Sigma, \phi)$ to derive updated estimates.

According to the standard theory, this leads to maximisation of the (incomplete) data log-likelihood

The full log-likelihood given the country-specific means $\beta$ are known, is of the form:

$$l(\mu, \Sigma, \phi) = -\frac{n}{2}\log(2\pi) - \frac{1}{2}\log(|\Sigma|) - \frac{1}{2}(\beta - \mu)^T \Sigma^{-1} (\beta - \mu)$$

$$-\frac{N}{2}\log(2\pi) - Nlog(\tau) - \sum_{j=1}^{N} (y_j - \phi^T z_j - \beta_{g(j)})^2 / 2\tau^2$$

Given the observed data, the country-specific means $\beta$ are given by:

$$\underline{b} = E\left(\underline{\beta}\middle|y\right) = \left(\Sigma^{-1} + diag(\frac{N_k}{\tau^2})\right)^{-1} \left(\Sigma^{-1}\mu + (\frac{\sum_{g(i)=k}(y_i - \phi^T z_i)}{\tau^2})\right)$$

$$= (\Sigma^{-1} + T^{-1}))^{-1}(\Sigma^{-1}\mu + T^{-1}Y'))$$

Where $Y = diag\left(\frac{1}{N_k}\sum_{g(i)=k}(y_i - \phi^T z_i)\right)$ is the mean of the PRS values for group *k*, adjusted for the covariates and $T = \tau^2 / diag(N_k)$

And variance-covariance matrix $S = (\Sigma^{-1} + T^{-1})^{-1}$

Where $N_k$is the number of individuals in country k.

The expectation of the first term in $l(\mu, \Sigma, \phi)$ is then (as in Clayton and Kaldor)

$$= -\frac{n}{2}\log(2\pi) - \frac{1}{2}\log(|\Sigma|) - \frac{1}{2}(b - \mu)^T \Sigma^{-1}(b - \mu) - \frac{1}{2}\sum_{i,j}\Sigma_{ij}^{-1} S_{ij}$$

The second part is similar:

$$-\frac{N}{2}\log(2\pi) - Nlog(\tau) - \sum_{j=1}^{N} \frac{\left(y_j - \phi^T z_j - b_{g(j)}\right)^2}{2\tau^2} - \frac{1}{2\tau^2}\sum_{k} N_k S_{kk}$$

Where $n$ is the total number of countries, $N = \sum N_k$ is the total number of individuals

$$Q(\mu, \Sigma, \phi) = -\frac{n}{2}\log(2\pi) - \frac{1}{2}\log(|\Sigma|) - \frac{1}{2}(b-\mu)^T\Sigma^{-1}(b-\mu) - \frac{1}{2}\sum_{i,j}\Sigma_{ij}^{-1}S_{ij} - \frac{N}{2}\log(2\pi) - Nlog(\tau) - \sum_{j=1}^{N}\frac{\left(y_j - \phi^T z_j - b_{g(j)}\right)^2}{2\tau^2} - \frac{1}{2\tau^2}\sum_k N_k S_{jj}$$

Following Clayton and Kaldor, we replace $D = \sigma^2\Sigma^{-1}$

$$Q(\mu, \Sigma, \phi) = -\frac{n}{2}\log(2\pi) + \frac{1}{2}\log(|\mathrm{D}|) - n\,log(\sigma) - \frac{1}{2\sigma^2}\left((b-\mu)^T D(b-\mu) + \sum_{i,j} D_{ij}S_{ij}\right) - \frac{N}{2}\log(2\pi) - Nlog(\tau) - \sum_{j=1}^{N}\frac{\left(y_j - \phi^T z_j - b_{g(j)}\right)^2}{2\tau^2} - \frac{1}{2\tau^2}\sum_k N_k S_{kk}$$

$Q(\mu, \Sigma, \phi)$ is the sum of two components, which can be maximised separately in the M-Step. The first part depends on $\mu$ and $\sigma^2$, while the second part is a simple linear regression log-likelihood, which can be maximised, to estimate $\phi$and $\tau^2$ [once the country-specific means b are considered known, the first part doesn't depend on $\phi$]. The first part maximises to give:

$$\hat{\mu} = \left(\sum D_{ij}\right)^{-1}\left(\sum D_{ij}b_j\right)$$

$$\widehat{\sigma^2} = \frac{1}{n}\left((b-\mu)^T D(b-\mu) + \sum_{i,j} D_{ij}S_{ij}\right)$$

Substitution back into $Q(\mu, \Sigma, \phi)$ gives

$$\mathrm{g(D)} = \mathrm{Constant} + \log(|\mathrm{D}|) - n\,log(\sigma^2) - 2Nlog(\tau) - \sum_{j=1}^{N}\frac{(y_j - y')^2}{\tau^2} = \mathrm{Constant} + \log(|\mathrm{D}|) - n\,log(\sigma^2)$$

To be maximised over D, as in Clayton & Kaldor.

Clayton and Kaldor, consider correlations between countries such that the covariance matrix $\Sigma$ is assumed to be of the form: $\Sigma = \sigma^2(1-\rho W)^{-1}$ where $W_{ij} = 1$ if countries i and j are neighbouring and 0 otherwise. $\rho$ is an autocorrelation parameter (0 implies country-specific estimates are uncorrelated). Other symmetric matrices can also be considered.

Using this adjacency matrix, this comes down to maximising g(D) over values of the parameter $\rho$. This is most easily accomplished using a grid search for $\rho$ over the interval $(0, \frac{1}{\lambda_{max}})$ where$\lambda_{max}$ is the largest eigenvalue of W (the maximum possible value of $\rho$ is determined by the requirement that $D = I - \rho W$ must be positive definite for $\Sigma$ to be a covariance matrix. Therefore, the solutions to $|\mathrm{D} - \lambda^* I| = 0$ , the eigenvalues of D, must all be positive, or equivalently:

$$\left|W - (\frac{1-\lambda^*}{\rho})I\right| = 0$$

So for all eigenvalues of W, $\lambda^* = 1 - \rho\lambda > 0$ so $\rho < 1/\lambda$ for all eigenvalues.

Note that $|\mathrm{D}| = \prod(1 - \lambda_k)$ where $\lambda_k$are the eigenvalues of W.

The second part of $Q(\mu, \Sigma, \phi)$ can maximised to obtain estimates of $\phi$ using a simple linear regression of the form (in R):

lm(y~z, offset(b) – 1) where the country-specific means are now considered fixed offsets. The corresponding estimate of $\tau^2$ is given by:

$$\widehat{\tau^2} = \frac{1}{N}\sum_{j=1}^{N}\left(y_j - \phi^T z_j - b_{g(j)}\right)^2 + \frac{1}{N}\sum_{k=1}^{n} N_k S_{kk}$$

Where the first term can be computed from the mean residuals.

### Supplementary Figures

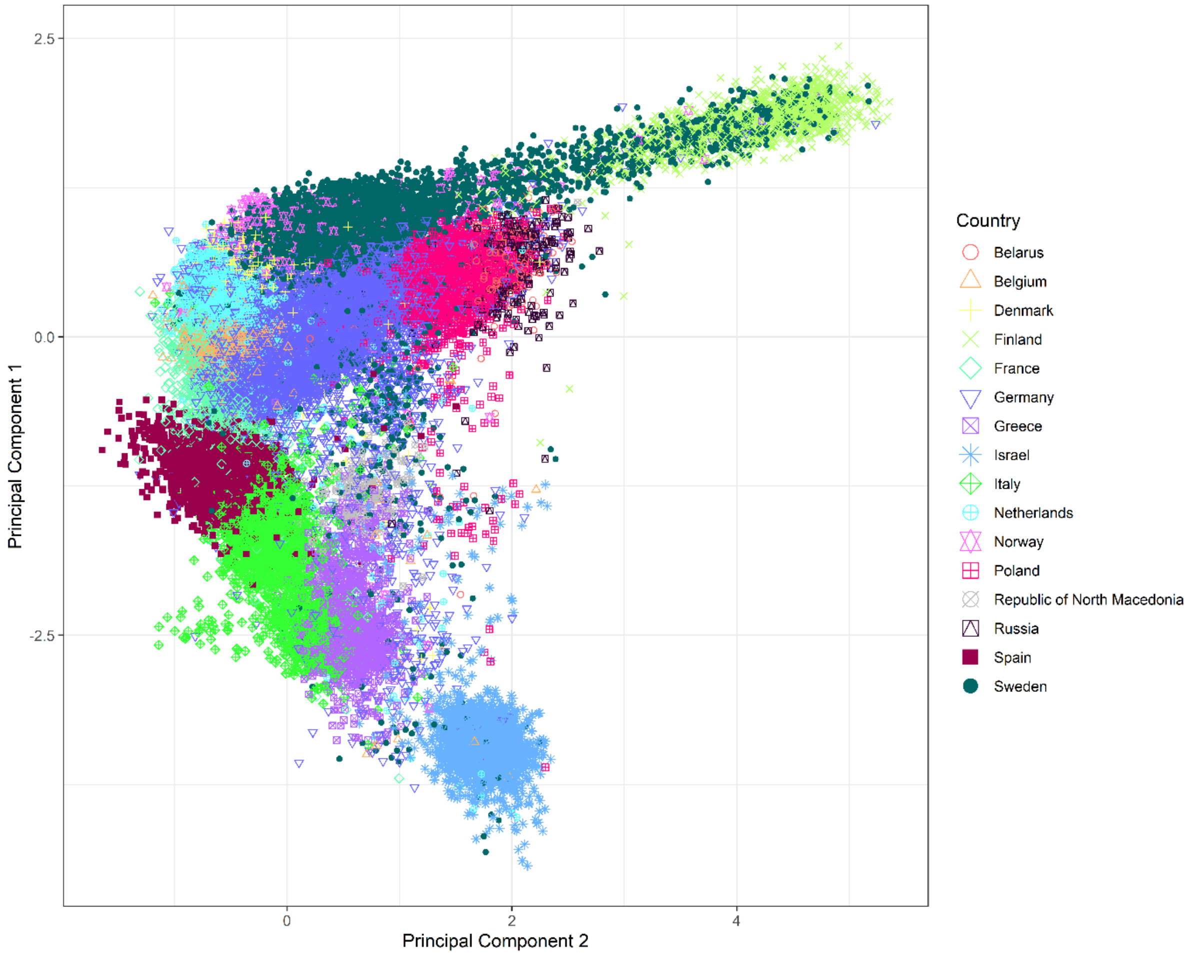


Supplementary Figure 1: Plot of the first two principal components for BCAC individuals genotyped with OncoArray. Individuals from the same country are plotted with the same colour and shape.

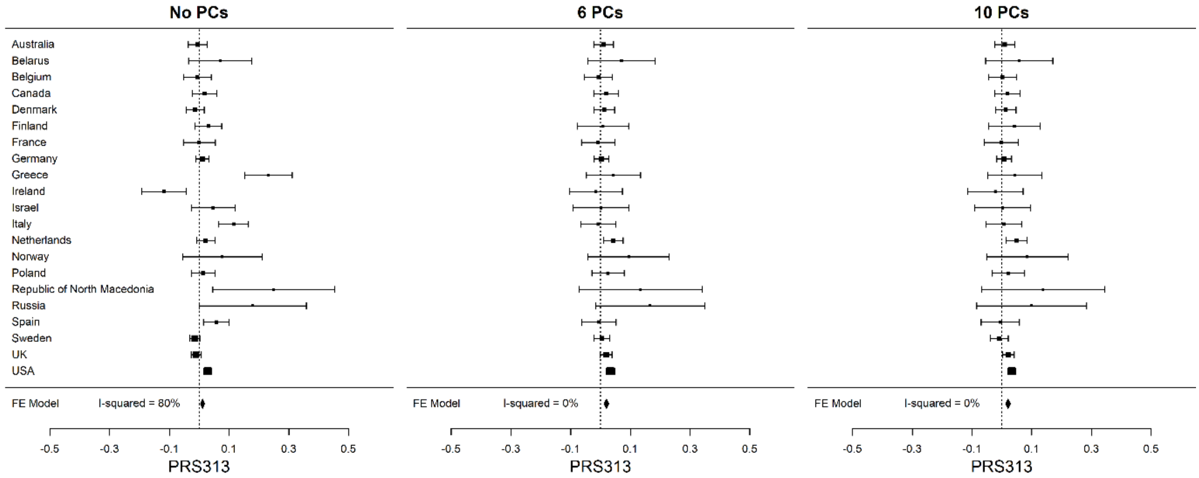


Supplementary Figure 2: Distribution of the mean $PRS_{313}$ for overall breast cancer across the countries in the control dataset of BCAC, without PC adjustment and when adjusted for the first 6 PCs and 10 PCs and array. The squares represent the mean PRS by country and the error bars represent the corresponding 95% confidence intervals (PCs: Principal components; FE Model: Fixed effect Model).

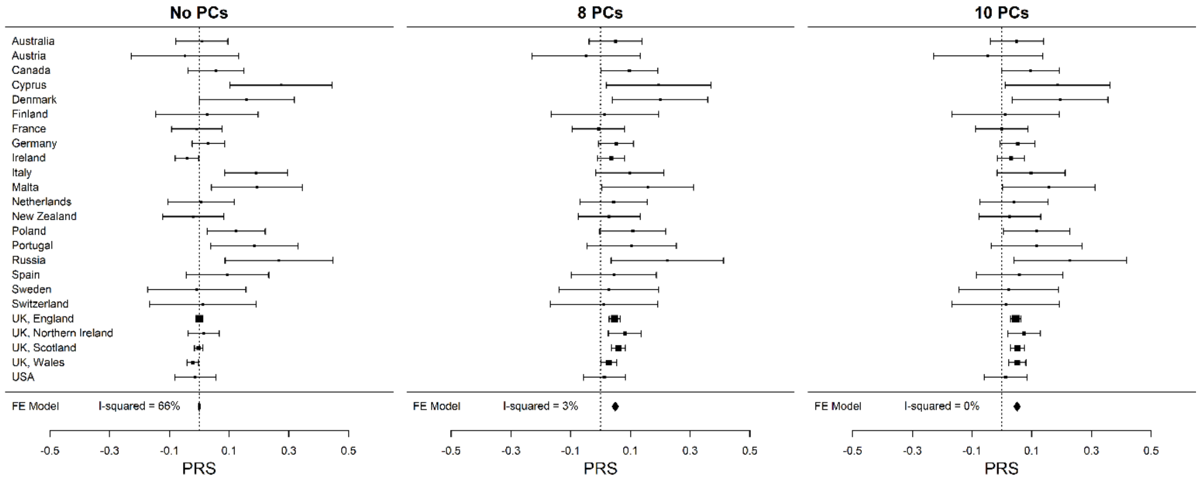


Supplementary Figure 3: Distribution of the mean $PRS_{306}$ for overall breast cancer across the countries in the white female individuals of the UK Biobank dataset, without PC adjustment and when adjusted for the first 8 PCs and 10 PCs. The squares represent the mean PRS by country and the error bars represent the corresponding 95% confidence intervals (PCs: Principal components; FE Model: Fixed effect Model).
