## Supplementary Tables for "Differences in polygenic score distributions in European ancestry populations: implications for breast cancer risk prediction"

**Supplementary Table 1A**: Studies participating in BCAC by country used in the analysis.

| Study | Controls | Cases | Total | Acronym | Study design |
|---|---|---|---|---|---|
| **Australia** | | | | | |
| ABCFS | 738 | 1432 | 2170 | Australian Breast Cancer Family Study | Case-control study |
| ABCTB | 375 | 920 | 1295 | Australian Breast Cancer Tissue Bank | Case-control study |
| BCEES | 835 | 783 | 1618 | Breast Cancer Employment and Environment Study | Case-control study |
| KCONFAB/AOCS | 896 | 462 | 1358 | Kathleen Cuningham Foundation Consortium for research into Familial Breast Cancer/Australian Ovarian Cancer Study | Case-control study |
| MCCS | 1205 | 1065 | 2270 | Melbourne Collaborative Cohort Study | Prospective cohort: nested case-control study |
| **Total** | 4049 | 4662 | 8711 | | |
| **Belarus** | | | | | |
| HMBCS | 342 | 744 | 1086 | Hannover-Minsk Breast Cancer Study | Case-control study |
| **Belgium** | | | | | |
| LMBC | 1823 | 3398 | 5221 | Leuven Multidisciplinary Breast Centre | Case-control study |
| **Canada** | | | | | |
| CBCS | 817 | 568 | 1385 | Canadian Breast Cancer Study | Case-control study |
| MTLGEBCS | 464 | 533 | 997 | Montreal Gene-Environment Breast Cancer Study | Case-control study |
| OFBCR | 996 | 2209 | 3205 | Ontario Familial Breast Cancer Registry | Case-control study |
| **Total** | 2277 | 3310 | 5587 | | |
| **Denmark** | | | | | |
| CGPS | 5241 | 4275 | 9516 | Copenhagen General Population Study | Case-control study |
| **Finland** | | | | | |
| HEBCS | 1236 | 1796 | 3032 | Helsinki Breast Cancer Study | Case-control study |
| KBCP | 433 | 543 | 976 | Kuopio Breast Cancer Project | Case-control study |
| OBCS | 414 | 499 | 913 | Oulu Breast Cancer Study | Case-control study |
| **Total** | 2083 | 2838 | 4921 | | |
| **France** | | | | | |
| CECILE | 1002 | 910 | 1912 | CECILE Breast Cancer Study | Case-control study |
| EPIC | 370 | 378 | 748 | European Prospective Investigation Into Cancer and Nutrition | Prospective cohort: nested case-control study |
| **Total** | 1372 | 1288 | 2660 | | |
| **Germany** | | | | | |
| BBCC | 706 | 839 | 1545 | Bavarian Breast Cancer Cases and Controls | Case-control study |

| | | | | | |
|---|---|---|---|---|---|
| BSUCH | 1119 | 991 | 2110 | Breast Cancer Study of the University of Heidelberg | Case-control study |
| EPIC | 650 | 586 | 1236 | European Prospective Investigation Into Cancer and Nutrition | Prospective cohort: nested case-control study |
| ESTHER | 505 | 475 | 980 | ESTHER Breast Cancer Study | Case-control study |
| GC-HBOC | 1732 | 3378 | 5110 | German Consortium for Hereditary Breast & Ovarian Cancer | Case-control study |
| GENICA | 710 | 912 | 1622 | Gene Environment Interaction and Breast Cancer in Germany | Case-control study |
| GEPARSIXTO | 0 | 387 | 387 | Randomized phase II trial | Case-only study |
| GESBC | 181 | 312 | 493 | Genetic Epidemiology Study of Breast Cancer by Age 50 | Case-control study |
| HABCS | 866 | 909 | 1775 | Hannover Breast Cancer Study | Case-control study |
| MARIE | 2065 | 1642 | 3707 | Mammary Carcinoma Risk Factor Investigation | Case-control study |
| PREFACE | 0 | 2954 | 2954 | Evaluation of Predictive Factors regarding the Effectivity of Aromatase Inhibitor Therapy | Case-only study |
| SKKDKFZS | 29 | 1220 | 1249 | Städtisches Klinikum Karlsruhe Deutsches Krebsforschungszentrum Study | Case-only study |
| SUCCESSB | 0 | 440 | 440 | Simultaneous Study of Gemcitabine-Docetaxel Combination adjuvant treatment | Case-only study |
| SUCCESSC | 0 | 2836 | 2836 | Simultaneous Study of Docetaxel Based Anthracycline Free Adjuvant Treatment Evaluation | Case-only study |
| **Total** | 8563 | 17881 | 26444 | | |
| **Greece** | | | | | |
| CCGP | 332 | 667 | 999 | Crete Cancer Genetics Program | Case-control study |
| EPIC | 180 | 173 | 353 | European Prospective Investigation Into Cancer and Nutrition | Prospective cohort: nested case-control study |
| TNBCC | 95 | 412 | 507 | Triple-Negative Breast Cancer Consortium | Case-control studies |
| **Total** | 607 | 1252 | 1859 | | |
| **Ireland** | | | | | |
| BIGGS | 719 | 793 | 1512 | Breast Cancer in Galway Genetic Study | Case-control study |
| **Israel** | | | | | |
| BCINIS | 724 | 1337 | 2061 | Breast Cancer in Northern Israel Study | Case-control study |
| **Italy** | | | | | |
| EPIC | 788 | 743 | 1531 | European Prospective Investigation Into Cancer and Nutrition | Prospective cohort: nested case-control study |
| MBCSG | 766 | 738 | 1504 | Milan Breast Cancer Study Group | Case-control study |
| **Total** | 1554 | 1481 | 3035 | | |
| **Netherlands** | | | | | |
| ABCS | 1817 | 1129 | 2946 | Amsterdam Breast Cancer Study | Case-control study |
| EPIC | 676 | 630 | 1306 | European Prospective Investigation Into Cancer and Nutrition | Prospective cohort: nested case-control study |
| ORIGO | 986 | 1239 | 2225 | Leiden University Medical Centre Breast Cancer Study | Prospective cohort: nested case-control study |
| RBCS | 928 | 1029 | 1957 | Rotterdam Breast Cancer Study | Case-control study |
| ABCS-F | 0 | 989 | 989 | Amsterdam Breast Cancer Study – Familial | Case-only study |
| **Total** | 4407 | 5016 | 9423 | | |
| **Norway** | | | | | |
| NBCS | 217 | 2386 | 2603 | Norwegian Breast Cancer Study | Case-control study |
| **Poland** | | | | | |

| | | | | | |
|---|---|---|---|---|---|
| PBCS | 2082 | 1767 | 3849 | NCI Polish Breast Cancer Study | Case-control study |
| SZBCS | 472 | 683 | 1155 | IHCC-Szczecin Breast Cancer Study | Case-control study |
| **Total** | 2554 | 2450 | 5004 | | |
| **Republic of North Macedonia** |  |  |  |  |  |
| MABCS | 92 | 78 | 170 | Macedonian Breast Cancer Study | Case-control study |
| **Russia** | | | | | |
| HUBCS | 120 | 211 | 331 | Hannover-Ufa Breast Cancer Study | Case-control study |
| **Spain** | | | | | |
| BREOGAN | 916 | 1536 | 2452 | Breast Oncology Galicia Network | Case-control study |
| CNIO-BCS | 871 | 866 | 1737 | Spanish National Cancer Centre Breast Cancer Study | Case-control study |
| EPIC | 311 | 299 | 610 | European Prospective Investigation Into Cancer and Nutrition | Prospective cohort: nested case-control study |
| HCSC | 0 | 423 | 423 | Hospital Clinico San Carlos | Case-control study |
| **Total** | 2098 | 3124 | 5222 | | |
| **Sweden** | | | | | |
| KARBAC | 658 | 806 | 1464 | Karolinska Breast Cancer Study | Case-control study |
| KARMA | 6983 | 2839 | 9822 | Karolinska Mammography Project for Risk Prediction of Breast Cancer – Cohort Study | Case-control study |
| MISS | 1545 | 633 | 2178 | Melanoma Inquiry of Southern Sweden | Prospective cohort: nested case-control study |
| PKARMA | 5417 | 5041 | 10458 | Karolinska Mammography Project for Risk Prediction of Breast Cancer - Case-Control Study | Case-control study |
| SASBAC | 1373 | 1129 | 2502 | Singapore and Sweden Breast Cancer Study | Case-control study |
| SMC | 704 | 1509 | 2213 | Swedish Mammography Cohort | Prospective cohort: nested case-control study |
| **Total** | 16680 | 11957 | 28637 | | |
| **UK** | | | | | |
| BBCS | 1838 | 1525 | 3363 | British Breast Cancer Study | Case-control study |
| DIETCOMPLYF | 0 | 708 | 708 | DietCompLyf Breast Cancer Survival Study | Prospective cohort: nested case-control study |
| EPIC | 669 | 626 | 1295 | European Prospective Investigation Into Cancer and Nutrition | Prospective cohort: nested case-control study |
| FHRISK | 943 | 146 | 1089 | Family History Risk Study | Case-control study |
| GLACIER | 0 | 1918 | 1918 | Study to Investigate the Genetics of Lobular Carcinoma In situ in Europe | |
| ICICLE | 1 | 204 | 205 | Study to Investigate the Genetics of In Situ Carcinoma of the Ductal Subtype | |
| POSH | 0 | 1088 | 1088 | Prospective Study of Outcomes in Sporadic Versus Hereditary Breast Cancer | Case-only study |
| PROCAS | 1648 | 380 | 2028 | Predicting the Risk Of Cancer At Screening Study | Prospective cohort: nested case-control study |
| SBCS | 848 | 878 | 1726 | Sheffield Breast Cancer Study | Case-control study |
| SEARCH | 8901 | 12805 | 21706 | Study of Epidemiology and Risk factors in Cancer Heredity | Case-control study |
| UKBGS | 1032 | 1053 | 2085 | UK Breakthrough Generations Study | Prospective cohort: nested case-control study |
| UKOPS | 974 | 0 | 974 | UK Ovarian Cancer Population Study | Case-control study |
| **Total** | 16854 | 21331 | 38185 | | |
| **USA** | | | | | |
| 2SISTER | 0 | 919 | 919 | The Two Sister Study | Case-only study |
| AHS | 1137 | 513 | 1650 | Agricultural Health Study | Prospective cohorts: nested case-control studies |
| BCFR-NY | 27 | 384 | 411 | New York site of the Breast Cancer Family Registry | Case-control study |

| | | | | | |
|---|---|---|---|---|---|
| BCFR-PA | 0 | 67 | 67 | Philadelphia site of the Breast Cancer Family Registry | Case-control study |
| BCFR-UTAH | 0 | 100 | 100 | Utah site of the Breast Cancer Family Registry | Case-control study |
| CPSII | 3315 | 2525 | 5840 | Cancer Prevention Study-II Nutrition Cohort | Prospective cohort: nested case-control study |
| CTS | 647 | 1176 | 1823 | California Teachers Study | Prospective cohort: nested case-control study |
| MCBCS | 2050 | 2071 | 4121 | Mayo Clinic Breast Cancer Study | Case-control study |
| MEC | 852 | 773 | 1625 | Multiethnic Cohort | Prospective cohort: nested case-control study |
| MMHS | 1635 | 275 | 1910 | Mayo Mammography Health Study | Prospective cohort: nested case-control study |
| MSKCC | 0 | 136 | 136 | Memorial Sloan-Kettering Cancer Center Study | Case-control study |
| NBHS | 731 | 572 | 1303 | Nashville Breast Health Study | Case-control study |
| NC-BCFR | 151 | 759 | 910 | Northern California Breast Cancer Family Registry | Case-control study |
| NCBCS | 1006 | 2074 | 3080 | North Carolina Breast Cancer study | Case-control study |
| NHS | 1804 | 1103 | 2907 | Nurses Health Study | Prospective cohort: nested case-control study |
| NHS2 | 1905 | 1112 | 3017 | Nurses Health Study 2 | Prospective cohort: nested case-control study |
| PLCO | 2595 | 1822 | 4417 | The Prostate, Lung, Colorectal and Ovarian (PLCO) Cancer Screening Trial | Prospective cohort: nested case-control study |
| SISTER | 1556 | 1504 | 3060 | The Sister Study | Prospective cohort: nested case-control study |
| TNBCC | 328 | 176 | 504 | Triple-Negative Breast Cancer Consortium | Case-control studies |
| UBCS | 0 | 606 | 606 | Utah Breast Cancer Study | |
| UCIBCS | 258 | 427 | 685 | UCI Breast Cancer Study | Case-control study |
| USRT | 1699 | 1354 | 3053 | US Radiologic Technologists Study | Case-control study |
| **Total** | 21696 | 20448 | 42144 | | |
| **Total** | **94072** | 110260 | 204332 | | |

**Supplementary Table 1B**: Total number of white female individuals participating in the UK Biobank stratified by country of birth, used in the analysis.

| Country | Controls used to calculate $PRS_{306}$ | Controls used to calculate standard PRS |
|---|---|---|
| Australia | 507 | 507 |
| Austria | 118 | 118 |
| Canada | 434 | 434 |
| Cyprus | 131 | 131 |
| Denmark | 152 | 152 |
| Finland | 130 | 130 |
| France | 539 | 538 |
| Germany | 1283 | 1281 |
| Ireland | 2462 | 2459 |
| Italy | 347 | 347 |
| Malta | 165 | 165 |
| Netherlands | 308 | 308 |
| New Zealand | 369 | 368 |
| Poland | 406 | 406 |
| Portugal | 180 | 180 |
| Russia | 118 | 118 |
| Spain | 201 | 201 |
| Sweden | 141 | 141 |
| Switzerland | 121 | 121 |
| UK, England | 184385 | 184106 |
| UK, Northern Ireland | 1454 | 1453 |
| UK, Scotland | 19658 | 19628 |
| UK, Wales | 10679 | 10668 |
| USA | 817 | 816 |
| **Total** | 225105 | 224776 |

**Supplementary Table 2**: The 313 variants included in the $PRS_{313}$ as calculated by Mavaddat et al., 2019 AJHG (1). Seven variants (indicated with an asterisk) of $PRS_{313}$ were not available in the UK Biobank.

| SNP name | Chromosome | Position b37[1] | Reference/Effect Allele | Effect size for Overall PRS[2] | Effect size for ER-positive PRS[3] | Effect size for ER-negative PRS[4] |
|---|---|---|---|---|---|---|
| 1_7917076_G_A | 1 | 7917076 | G/A | -0.0409 | -0.0393 | -0.0566 |
| 1_10566215_A_G | 1 | 10566215 | A/G | -0.0586 | -0.0407 | -0.1109 |
| 1_18807339_T_C | 1 | 18807339 | T/C | -0.0564 | -0.0649 | -0.0248 |
| 1_41380440_C_T | 1 | 41380440 | C/T | 0.0426 | 0.0423 | 0.0395 |
| 1_41389220_T_C | 1 | 41389220 | T/C | 0.1550 | 0.1377 | 0.1483 |
| 1_46670206_TC_T | 1 | 46670206 | TC/T | 0.0447 | 0.0595 | 0.0216 |
| 1_51467096_CT_C | 1 | 51467096 | CT/C | 0.0374 | 0.0426 | 0.0038 |
| 1_88156923_G_A | 1 | 88156923 | G/A | 0.0494 | 0.058 | 0.0183 |
| 1_88428199_C_A | 1 | 88428199 | C/A | -0.0387 | -0.0385 | -0.047 |
| 1_100880328_A_T | 1 | 100880328 | A/T | 0.0373 | 0.0355 | 0.016 |
| 1_110198129_CAAA_C | 1 | 110198129 | CAAA/C | 0.0458 | 0.0545 | 0.0266 |
| 1_114445880_G_A | 1 | 114445880 | G/A | 0.0621 | 0.0642 | 0.0579 |
| 1_118141492_A_C | 1 | 118141492 | A/C | 0.0452 | 0.0417 | 0.0551 |
| 1_120257110_T_C | 1 | 120257110 | T/C | 0.0385 | 0.043 | 0.0226 |
| 1_121280613_A_G | 1 | 121280613 | A/G | 0.0881 | 0.1052 | 0.0209 |
| 1_121287994_A_G | 1 | 121287994 | A/G | -0.0673 | -0.0814 | -0.0114 |
| 1_145604302_C_CT | 1 | 145604302 | C/CT | -0.0399 | -0.0469 | -0.0126 |
| 1_149906413_T_C | 1 | 149906413 | T/C | 0.0548 | 0.0625 | 0.0388 |
| 1_155556971_G_A | 1 | 155556971 | G/A | 0.0499 | 0.0606 | 0.0263 |
| 1_168171052_CA_C | 1 | 168171052 | CA/C | -0.0680 | -0.0774 | -0.0718 |
| 1_172328767_T_TA | 1 | 172328767 | T/TA | -0.0435 | -0.0417 | -0.0655 |
| 1_201437832_C_T | 1 | 201437832 | C/T | 0.0917 | 0.0815 | 0.0714 |
| 1_202184600_C_T | 1 | 202184600 | C/T | -0.0065 | 0.0133 | -0.0822 |
| 1_203770448_T_A | 1 | 203770448 | T/A | 0.0498 | 0.0472 | 0.0332 |
| 1_204502514_T_TTCTGAAACAGGG | 1 | 204502514 | T/TTCTGAAACAGGG | -0.0321 | -0.0024 | -0.1345 |
| 1_208076291_G_A | 1 | 208076291 | G/A | -0.0366 | -0.0313 | -0.0278 |
| 1_217053815_T_G | 1 | 217053815 | T/G | 0.0417 | 0.0409 | 0.0472 |
| 1_217220574_G_A | 1 | 217220574 | G/A | -0.0440 | -0.0459 | 0.0029 |
| 1_220671050_C_T | 1 | 220671050 | C/T | 0.0418 | 0.0482 | 0.0153 |
| 1_242034263_A_G | 1 | 242034263 | A/G | 0.1428 | 0.1519 | 0.1413 |
| 2_10138983_T_C | 2 | 10138983 | T/C | 0.0603 | 0.0596 | 0.0458 |
| 2_19315675_T_A | 2 | 19315675 | T/A | -0.0331 | -0.0229 | -0.057 |
| 2_25129473_A_G | 2 | 25129473 | A/G | -0.0427 | -0.0365 | -0.067 |
| 2_29179452_G_C | 2 | 29179452 | G/C | -0.0066 | 0.0207 | -0.1006 |
| 2_29615233_T_C | 2 | 29615233 | T/C | -0.0427 | -0.0489 | -0.0371 |
| 2_39699510_C_CT | 2 | 39699510 | C/CT | -0.0402 | -0.0336 | -0.0538 |
| 2_70172587_G_A | 2 | 70172587 | G/A | -0.0412 | -0.0334 | -0.0361 |
| 2_88358825_G_C | 2 | 88358825 | G/C | 0.0473 | 0.0443 | 0.0505 |
| 2_121058254_A_G | 2 | 121058254 | A/G | -0.0334 | -0.0232 | -0.0682 |
| 2_121089731_T_C | 2 | 121089731 | T/C | -0.0427 | -0.029 | -0.1027 |
| 2_121159205_G_A | 2 | 121159205 | G/A | -0.0440 | -0.0507 | -0.0162 |
| 2_121246568_T_C | 2 | 121246568 | T/C | 0.0992 | 0.092 | 0.1112 |
| 2_172974566_C_G | 2 | 172974566 | C/G | -0.0473 | -0.0611 | -0.0061 |
| 2_174212910_A_G | 2 | 174212910 | A/G | 0.0593 | 0.0621 | 0.0175 |
| 2_192381934_C_T | 2 | 192381934 | C/T | 0.0316 | 0.018 | 0.1012 |
| 2_202204741_T_C | 2 | 202204741 | T/C | -0.0492 | -0.0505 | -0.0526 |
| 2_217920769_G_T | 2 | 217920769 | G/T | -0.1318 | -0.1532 | -0.0589 |
| 2_217955896_GA_G | 2 | 217955896 | GA/G | -0.2016 | -0.2362 | -0.0558 |
| 2_218292158_C_G | 2 | 218292158 | C/G | -0.0757 | -0.0813 | -0.0599 |
| 2_218714845_G_A | 2 | 218714845 | G/A | -0.0431 | -0.0463 | -0.0184 |
| 2_241388857_C_A | 2 | 241388857 | C/A | -0.1232 | -0.1335 | -0.1727 |
| 3_4742251_A_G | 3 | 4742251 | A/G | 0.0616 | 0.0609 | 0.0422 |
| 3_27353716_C_A | 3 | 27353716 | C/A | 0.0748 | 0.0822 | 0.031 |
| 3_27388664_C_G | 3 | 27388664 | C/G | 0.0502 | 0.0539 | 0.0297 |
| 3_29294845_C_T | 3 | 29294845 | C/T | -0.1281 | -0.1221 | -0.2988 |
| 3_30684907_C_T | 3 | 30684907 | C/T | 0.0592 | 0.0657 | 0.017 |
| 3_46888198_T_C | 3 | 46888198 | T/C | -0.0806 | -0.0842 | -0.0716 |
| 3_49709912_C_CT | 3 | 49709912 | C/CT | -0.0367 | -0.0355 | -0.0721 |
| 3_55970777_A_AT | 3 | 55970777 | A/AT | -0.1195 | -0.124 | -0.0586 |
| 3_59373745_C_T | 3 | 59373745 | C/T | -0.0394 | -0.0439 | -0.0398 |
| 3_63887449_T_TTG* | 3 | 63887449 | T/TTG | 0.0648 | 0.0627 | 0.043 |
| 3_71620370_T_G | 3 | 71620370 | T/G | -0.0374 | -0.0345 | -0.0379 |
| 3_87037543_A_G | 3 | 87037543 | A/G | -0.0723 | -0.0726 | -0.0531 |
| 3_99403877_G_A | 3 | 99403877 | G/A | -0.0376 | -0.0378 | -0.0279 |
| 3_141112859_CTT_C | 3 | 141112859 | CTT/C | 0.0551 | 0.0607 | 0.0282 |
| 3_172285237_G_A | 3 | 172285237 | G/A | 0.0422 | 0.0501 | -0.0133 |
| 3_189774456_C_T | 3 | 189774456 | C/T | -0.0478 | -0.0469 | -0.0449 |

| | | | | | | |
|---|---|---|---|---|---|---|
| 4_38784633_G_T | 4 | 38784633 | G/T | 0.0489 | 0.0495 | 0.0497 |
| 4_84370124_TAA_TA* | 4 | 84370124 | TAA/TA | -0.0464 | -0.0438 | -0.0489 |
| 4_89240476_G_A | 4 | 89240476 | G/A | 0.0352 | 0.0392 | 0.0261 |
| 4_92594859_TTCTTTC_T | 4 | 92594859 | TTCTTTC/T | -0.0407 | -0.0377 | -0.0359 |
| 4_106069013_G_T | 4 | 106069013 | G/T | 0.0471 | 0.0594 | 0.0097 |
| 4_126752992_A_AAT* | 4 | 126752992 | A/AAT | -0.0377 | -0.0361 | -0.0638 |
| 4_143467195_C_T | 4 | 143467195 | C/T | -0.0569 | -0.0613 | -0.0594 |
| 4_151218296_CATATTT_C | 4 | 151218296 | CATATTT/C | 0.0388 | 0.0307 | 0.0557 |
| 4_175842495_G_A | 4 | 175842495 | G/A | -0.0898 | -0.1162 | 0.0199 |
| 4_175847436_C_A | 4 | 175847436 | C/A | 0.0348 | 0.0537 | -0.0099 |
| 4_187503758_A_T* | 4 | 187503758 | A/T | 0.0357 | 0.0352 | 0.0194 |
| 5_345109_T_C | 5 | 345109 | T/C | 0.0840 | 0.0856 | 0.0681 |
| 5_1279790_C_T | 5 | 1279790 | C/T | 0.0617 | 0.0325 | 0.1502 |
| 5_1296255_A_AG | 5 | 1296255 | A/AG | -0.0549 | -0.0417 | -0.1056 |
| 5_1353077_T_C | 5 | 1353077 | T/C | 0.1552 | 0.1572 | 0.1214 |
| 5_2777029_G_A | 5 | 2777029 | G/A | 0.0391 | 0.041 | 0.0231 |
| 5_16231194_G_C | 5 | 16231194 | G/C | -0.0426 | -0.0458 | -0.0404 |
| 5_32579616_TCA_T | 5 | 32579616 | TCA/T | 0.0363 | 0.0394 | 0.0072 |
| 5_44508264_G_GT | 5 | 44508264 | G/GT | -0.1177 | -0.126 | -0.1053 |
| 5_44619502_A_G | 5 | 44619502 | A/G | -0.1101 | -0.1186 | -0.0959 |
| 5_44649944_C_T | 5 | 44649944 | C/T | 0.0492 | 0.0713 | -0.0261 |
| 5_44706498_A_G | 5 | 44706498 | A/G | 0.0497 | 0.0648 | -0.0256 |
| 5_44853593_G_C | 5 | 44853593 | G/C | -0.0336 | -0.0222 | -0.0778 |
| 5_52679539_C_CA* | 5 | 52679539 | C/CA | 0.0571 | 0.0663 | 0.042 |
| 5_55662540_C_CT | 5 | 55662540 | C/CT | -0.0458 | -0.045 | -0.0299 |
| 5_55965167_C_T | 5 | 55965167 | C/T | 0.0394 | 0.0379 | 0.0405 |
| 5_56023083_T_G | 5 | 56023083 | T/G | 0.1366 | 0.1612 | 0.0686 |
| 5_56042972_C_T | 5 | 56042972 | C/T | 0.0865 | 0.1082 | 0.0058 |
| 5_56045081_T_C | 5 | 56045081 | T/C | -0.0564 | -0.0643 | -0.0168 |
| 5_58241712_C_T | 5 | 58241712 | C/T | -0.0434 | -0.0369 | -0.0408 |
| 5_71965007_G_A | 5 | 71965007 | G/A | -0.0410 | -0.0445 | -0.0238 |
| 5_73234583_T_C | 5 | 73234583 | T/C | -0.0363 | -0.0494 | -0.0101 |
| 5_77155397_GT_G | 5 | 77155397 | GT/G | -0.0408 | -0.0418 | -0.0489 |
| 5_79180995_G_GA | 5 | 79180995 | G/GA | 0.0328 | 0.0248 | 0.0804 |
| 5_81512947_TA_T | 5 | 81512947 | TA/T | -0.0598 | -0.0731 | -0.0342 |
| 5_90789470_G_A | 5 | 90789470 | G/A | -0.0564 | -0.0714 | -0.0031 |
| 5_104300273_G_T | 5 | 104300273 | G/T | -0.0487 | -0.0524 | -0.0271 |
| 5_122478676_C_A | 5 | 122478676 | C/A | -0.0386 | -0.0376 | -0.04 |
| 5_122705244_C_T | 5 | 122705244 | C/T | 0.0944 | 0.0963 | 0.0607 |
| 5_131640536_A_G | 5 | 131640536 | A/G | 0.0392 | 0.0467 | 0.0099 |
| 5_132407058_C_T | 5 | 132407058 | C/T | -0.0388 | -0.0561 | -0.0214 |
| 5_158244083_C_T | 5 | 158244083 | C/T | -0.0677 | -0.0635 | -0.0646 |
| 5_169591460_T_C | 5 | 169591460 | T/C | 0.0412 | 0.0501 | 0.0182 |
| 5_173358154_G_A | 5 | 173358154 | G/A | 0.0365 | 0.0395 | 0.0346 |
| 5_176134882_T_C | 5 | 176134882 | T/C | 0.0363 | 0.0368 | 0.0262 |
| 6_13713366_G_C | 6 | 13713366 | G/C | -0.0553 | -0.0623 | -0.0152 |
| 6_16399557_C_T | 6 | 16399557 | C/T | -0.0373 | -0.0435 | -0.0324 |
| 6_18783140_G_A | 6 | 18783140 | G/A | 0.0326 | 0.0478 | 0.0033 |
| 6_20537845_CA_C | 6 | 20537845 | CA/C | -0.0391 | -0.0416 | -0.0315 |
| 6_21923810_T_C | 6 | 21923810 | T/C | -0.0321 | -0.0438 | -0.0032 |
| 6_27425644_G_C | 6 | 27425644 | G/C | -0.0737 | -0.0838 | -0.0506 |
| 6_43227141_G_A | 6 | 43227141 | G/A | -0.0640 | -0.0614 | -0.0673 |
| 6_82263549_AAT_A | 6 | 82263549 | AAT/A | 0.0477 | 0.0406 | 0.0686 |
| 6_85912194_CAA_C | 6 | 85912194 | CAA/C | 0.0762 | 0.0569 | 0.0682 |
| 6_87803819_T_C | 6 | 87803819 | T/C | 0.0383 | 0.0318 | 0.0678 |
| 6_130341728_C_CT | 6 | 130341728 | C/CT | 0.0472 | 0.0433 | 0.0804 |
| 6_149595505_T_C | 6 | 149595505 | T/C | -0.0476 | -0.0601 | -0.0235 |
| 6_151949806_A_C | 6 | 151949806 | A/C | 0.0703 | 0.0541 | 0.1103 |
| 6_151955914_A_G | 6 | 151955914 | A/G | 0.1449 | 0.115 | 0.224 |
| 6_152022664_CAAAAAAA_C | 6 | 152022664 | CAAAAAAA/C | 0.0137 | 0.0185 | -0.017 |
| 6_152023191_G_A | 6 | 152023191 | G/A | 0.0626 | 0.0509 | 0.1008 |
| 6_152055978_A_T | 6 | 152055978 | A/T | 0.0740 | 0.0634 | 0.108 |
| 6_152432902_C_T | 6 | 152432902 | C/T | 0.0649 | 0.0527 | 0.0965 |
| 6_169006947_C_G | 6 | 169006947 | C/G | -0.0308 | -0.0252 | -0.0628 |
| 6_170332621_T_C | 6 | 170332621 | T/C | 0.0373 | 0.0403 | 0.0334 |
| 7_21940960_A_G | 7 | 21940960 | A/G | -0.0467 | -0.0413 | -0.0574 |
| 7_25569548_C_T | 7 | 25569548 | C/T | -0.0486 | -0.0485 | -0.0682 |
| 7_28869017_G_A | 7 | 28869017 | G/A | -0.0572 | -0.0504 | -0.0487 |
| 7_55192256_A_C | 7 | 55192256 | A/C | -0.0349 | -0.0269 | -0.0536 |
| 7_91459189_A_ATT* | 7 | 91459189 | A/ATT | 0.0452 | 0.0439 | 0.0486 |
| 7_94113799_T_C | 7 | 94113799 | T/C | 0.0449 | 0.0489 | 0.0116 |

| | | | | | | |
|---|---|---|---|---|---|---|
| 7_98005235_G_A | 7 | 98005235 | G/A | -0.0467 | -0.0466 | -0.0267 |
| 7_99948655_T_G | 7 | 99948655 | T/G | 0.0420 | 0.0385 | 0.0497 |
| 7_101552440_G_A | 7 | 101552440 | G/A | -0.0568 | -0.0742 | -0.0241 |
| 7_102481842_T_C | 7 | 102481842 | T/C | 0.0418 | 0.0406 | 0.0411 |
| 7_130656911_C_T | 7 | 130656911 | C/T | -0.0476 | -0.0522 | -0.025 |
| 7_130674481_G_A | 7 | 130674481 | G/A | 0.0416 | 0.0407 | 0.028 |
| 7_139943702_CT_C | 7 | 139943702 | CT/C | 0.0582 | 0.0666 | 0.0057 |
| 7_144048902_G_T | 7 | 144048902 | G/T | -0.0563 | -0.0592 | -0.0148 |
| 8_170692_T_C | 8 | 170692 | T/C | 0.0477 | 0.0348 | 0.104 |
| 8_17787610_CT_C | 8 | 17787610 | CT/C | -0.0377 | -0.0367 | -0.0295 |
| 8_23447496_A_G | 8 | 23447496 | A/G | -0.0389 | -0.0361 | -0.0426 |
| 8_23663653_C_A | 8 | 23663653 | C/A | 0.0335 | 0.0451 | 0.0059 |
| 8_29509616_A_C | 8 | 29509616 | A/C | -0.0601 | -0.0655 | -0.0512 |
| 8_36858483_A_G | 8 | 36858483 | A/G | -0.0760 | -0.0713 | -0.1013 |
| 8_76230943_A_G | 8 | 76230943 | A/G | 0.0755 | 0.0817 | 0.0617 |
| 8_76333056_C_T | 8 | 76333056 | C/T | 0.1129 | 0.1216 | 0.0879 |
| 8_76378165_G_T | 8 | 76378165 | G/T | -0.0391 | -0.0418 | -0.026 |
| 8_102483100_T_C | 8 | 102483100 | T/C | 0.0593 | 0.0736 | 0.0137 |
| 8_106358620_A_T | 8 | 106358620 | A/T | -0.0745 | -0.0895 | -0.01 |
| 8_117209548_A_G | 8 | 117209548 | A/G | -0.0417 | -0.0456 | -0.0409 |
| 8_120862186_A_G | 8 | 120862186 | A/G | 0.0527 | 0.0598 | 0.0472 |
| 8_124563705_T_C | 8 | 124563705 | T/C | 0.0477 | 0.0465 | 0.0503 |
| 8_124571581_G_A | 8 | 124571581 | G/A | 0.0340 | 0.0355 | 0.0388 |
| 8_124739913_T_G | 8 | 124739913 | T/G | 0.0466 | 0.0395 | 0.0706 |
| 8_128213561_C_CA | 8 | 128213561 | C/CA | -0.0430 | -0.0468 | -0.04 |
| 8_128370949_C_G | 8 | 128370949 | C/G | 0.0642 | 0.082 | 0.0076 |
| 8_128372172_A_G | 8 | 128372172 | A/G | 0.0597 | 0.0508 | 0.057 |
| 8_129199566_G_A | 8 | 129199566 | G/A | 0.0615 | 0.0643 | 0.0505 |
| 8_143669254_A_G | 8 | 143669254 | A/G | -0.0346 | -0.0518 | -0.022 |
| 9_6880263_A_G | 9 | 6880263 | A/G | 0.0348 | 0.0499 | -0.0078 |
| 9_21964882_CAAAA_C | 9 | 21964882 | CAAAA/C | 0.0550 | 0.0467 | 0.0576 |
| 9_22041998_C_G | 9 | 22041998 | C/G | 0.0289 | 0.0168 | 0.0906 |
| 9_36928288_T_C | 9 | 36928288 | T/C | 0.0249 | 0.0259 | 0.0631 |
| 9_87782211_T_C | 9 | 87782211 | T/C | 0.0361 | 0.0432 | 0.0218 |
| 9_98362587_T_C | 9 | 98362587 | T/C | 0.0576 | 0.0625 | 0.0828 |
| 9_110303808_TAA_T | 9 | 110303808 | TAA/T | 0.0797 | 0.1007 | 0.013 |
| 9_110837073_A_G | 9 | 110837073 | A/G | 0.1158 | 0.1315 | 0.0289 |
| 9_110837176_C_T | 9 | 110837176 | C/T | 0.0653 | 0.0809 | -0.0037 |
| 9_110849525_G_T | 9 | 110849525 | G/T | 0.0153 | 0.0111 | 0.0336 |
| 9_110885479_C_T | 9 | 110885479 | C/T | 0.0877 | 0.111 | 0.0019 |
| 9_119313486_A_G | 9 | 119313486 | A/G | -0.0462 | -0.0477 | -0.0403 |
| 9_129424719_A_G | 9 | 129424719 | A/G | -0.0382 | -0.0437 | -0.0287 |
| 9_136146597_C_T | 9 | 136146597 | C/T | 0.0400 | 0.04 | 0.0253 |
| 10_5794652_A_G | 10 | 5794652 | A/G | 0.0470 | 0.0504 | 0.0385 |
| 10_13892298_G_A | 10 | 13892298 | G/A | 0.0371 | 0.0362 | 0.0382 |
| 10_22032942_A_G | 10 | 22032942 | A/G | -0.0580 | -0.0719 | 0.0344 |
| 10_22477776_ACC_A | 10 | 22477776 | ACC/A | 0.1687 | 0.1668 | 0.1849 |
| 10_22861490_A_C | 10 | 22861490 | A/C | 0.0875 | 0.096 | 0.0201 |
| 10_38523626_C_A | 10 | 38523626 | C/A | 0.0404 | 0.0381 | 0.0418 |
| 10_64299890_A_G | 10 | 64299890 | A/G | -0.1345 | -0.1428 | -0.103 |
| 10_64819996_G_T | 10 | 64819996 | G/T | 0.0472 | 0.0442 | 0.0403 |
| 10_71335574_C_T | 10 | 71335574 | C/T | -0.0404 | -0.0411 | -0.054 |
| 10_80851257_G_T | 10 | 80851257 | G/T | -0.0805 | -0.0898 | -0.0443 |
| 10_80886726_A_G | 10 | 80886726 | A/G | 0.0762 | 0.078 | 0.0444 |
| 10_95292187_CAA_C | 10 | 95292187 | CAA/C | -0.0512 | -0.049 | -0.0419 |
| 10_114777670_C_T | 10 | 114777670 | C/T | 0.0472 | 0.0422 | 0.0559 |
| 10_115128491_T_C | 10 | 115128491 | T/C | -0.0592 | -0.0602 | -0.0592 |
| 10_123095209_G_A | 10 | 123095209 | G/A | -0.0538 | -0.0702 | 0.0048 |
| 10_123340107_A_G | 10 | 123340107 | A/G | 0.1508 | 0.1837 | 0.0053 |
| 10_123340431_GC_G | 10 | 123340431 | GC/G | -0.2408 | -0.2913 | -0.0326 |
| 10_123349324_A_T | 10 | 123349324 | A/T | -0.2609 | -0.327 | -0.0137 |
| 11_433617_T_C | 11 | 433617 | T/C | -0.0437 | -0.0494 | -0.0322 |
| 11_803017_A_G | 11 | 803017 | A/G | 0.0457 | 0.04 | 0.0559 |
| 11_1895708_C_A | 11 | 1895708 | C/A | -0.0762 | -0.0788 | -0.0538 |
| 11_18664241_T_G | 11 | 18664241 | T/G | 0.0461 | 0.0455 | 0.0633 |
| 11_42844441_C_T | 11 | 42844441 | C/T | -0.0336 | -0.0334 | -0.0669 |
| 11_44368892_G_A | 11 | 44368892 | G/A | 0.0374 | 0.0357 | 0.021 |
| 11_46318032_C_G | 11 | 46318032 | C/G | -0.0748 | -0.0693 | -0.0688 |
| 11_65553492_C_A | 11 | 65553492 | C/A | 0.0425 | 0.0444 | 0.0305 |
| 11_65572431_G_A | 11 | 65572431 | G/A | -0.0347 | -0.0448 | -0.0067 |
| 11_69328130_A_T | 11 | 69328130 | A/T | -0.0423 | -0.0538 | 0.0143 |

| | | | | | | |
|---|---|---|---|---|---|---|
| 11_69330983_G_A | 11 | 69330983 | G/A | 0.1022 | 0.124 | 0.0174 |
| 11_69331418_C_T | 11 | 69331418 | C/T | 0.1782 | 0.2018 | 0.0066 |
| 11_103614438_T_G | 11 | 103614438 | T/G | 0.0147 | 0.0029 | 0.0676 |
| 11_108267402_C_CA | 11 | 108267402 | C/CA | -0.0022 | 0.0141 | -0.0629 |
| 11_111696440_T_C | 11 | 111696440 | T/C | -0.0396 | -0.0435 | -0.0122 |
| 11_116727936_A_T | 11 | 116727936 | A/T | -0.0423 | -0.0372 | -0.062 |
| 11_122966626_A_G | 11 | 122966626 | A/G | -0.0383 | -0.0372 | -0.0484 |
| 11_129243417_T_G | 11 | 129243417 | T/G | -0.0543 | -0.0477 | -0.0605 |
| 11_129461016_A_G | 11 | 129461016 | A/G | 0.0453 | 0.0401 | 0.0594 |
| 12_293626_A_G | 12 | 293626 | A/G | 0.0401 | 0.0371 | 0.039 |
| 12_14413931_G_C | 12 | 14413931 | G/C | 0.0484 | 0.0411 | 0.054 |
| 12_28149568_C_T | 12 | 28149568 | C/T | -0.0620 | -0.0501 | -0.0683 |
| 12_28174817_C_T | 12 | 28174817 | C/T | -0.0856 | -0.083 | -0.101 |
| 12_28347382_C_T | 12 | 28347382 | C/T | -0.0521 | -0.0454 | -0.0469 |
| 12_29140260_G_A | 12 | 29140260 | G/A | 0.0647 | 0.069 | 0.0753 |
| 12_57146069_T_G | 12 | 57146069 | T/G | -0.0579 | -0.0585 | -0.0632 |
| 12_70798355_A_T | 12 | 70798355 | A/T | 0.0469 | 0.0471 | 0.015 |
| 12_83064195_G_GA | 12 | 83064195 | G/GA | 0.0671 | 0.0645 | 0.0717 |
| 12_85004551_C_T | 12 | 85004551 | C/T | 0.0348 | 0.0355 | 0.0358 |
| 12_96027759_A_G | 12 | 96027759 | A/G | -0.0867 | -0.0842 | -0.063 |
| 12_103097887_C_T | 12 | 103097887 | C/T | 0.0546 | 0.0611 | 0.0149 |
| 12_111600134_G_T | 12 | 111600134 | G/T | -0.0442 | -0.0441 | -0.0537 |
| 12_115108136_T_C | 12 | 115108136 | T/C | 0.0465 | 0.0533 | 0.0188 |
| 12_115796577_A_G | 12 | 115796577 | A/G | -0.0428 | -0.0643 | -0.0148 |
| 12_115835836_T_C | 12 | 115835836 | T/C | -0.0813 | -0.0977 | -0.0153 |
| 12_120832146_C_T | 12 | 120832146 | C/T | 0.0516 | 0.0534 | 0.0529 |
| 13_32839990_G_A | 13 | 32839990 | G/A | 0.0424 | 0.0386 | 0.0759 |
| 13_32972626_A_T | 13 | 32972626 | A/T | 0.2687 | 0.2308 | 0.4284 |
| 13_43501356_A_G | 13 | 43501356 | A/G | 0.0517 | 0.0458 | 0.0975 |
| 13_73806982_T_C | 13 | 73806982 | T/C | 0.0345 | 0.0251 | 0.0653 |
| 13_73960952_A_G | 13 | 73960952 | A/G | 0.0399 | 0.0368 | 0.073 |
| 14_37128564_C_A | 14 | 37128564 | C/A | -0.0733 | -0.085 | -0.0339 |
| 14_37228504_C_T | 14 | 37228504 | C/T | 0.0390 | 0.0408 | 0.0256 |
| 14_68660428_T_C | 14 | 68660428 | T/C | -0.0474 | -0.0612 | 0.0245 |
| 14_68979835_T_C | 14 | 68979835 | T/C | -0.0911 | -0.0972 | -0.0762 |
| 14_91751788_TC_T | 14 | 91751788 | TC/T | 0.0380 | 0.0447 | 0.0091 |
| 14_91841069_A_G | 14 | 91841069 | A/G | 0.0513 | 0.0553 | 0.0312 |
| 14_93070286_C_T | 14 | 93070286 | C/T | -0.0577 | -0.0519 | -0.0592 |
| 14_105213978_T_G | 14 | 105213978 | T/G | 0.0399 | 0.035 | 0.0403 |
| 15_46680811_C_A | 15 | 46680811 | C/A | -0.1973 | -0.1823 | -0.2337 |
| 15_50694306_A_G | 15 | 50694306 | A/G | -0.0417 | -0.0403 | -0.0392 |
| 15_66630569_G_A | 15 | 66630569 | G/A | -0.0369 | -0.0377 | -0.0343 |
| 15_67457698_A_G | 15 | 67457698 | A/G | 0.0782 | 0.099 | 0.0141 |
| 15_75750383_T_C | 15 | 75750383 | T/C | -0.0413 | -0.0419 | -0.0419 |
| 15_91512267_G_T | 15 | 91512267 | G/T | -0.0589 | -0.0557 | -0.0487 |
| 15_100905819_A_C | 15 | 100905819 | A/C | -0.0608 | -0.0599 | -0.0746 |
| 16_4008542_CAAAAA_C | 16 | 4008542 | CAAAAA/C | -0.0329 | -0.0184 | -0.0892 |
| 16_4106788_C_A | 16 | 4106788 | C/A | -0.0300 | -0.0182 | -0.0782 |
| 16_6963972_C_G | 16 | 6963972 | C/G | 0.0354 | 0.0303 | 0.0811 |
| 16_10706580_G_A | 16 | 10706580 | G/A | -0.0740 | -0.0763 | -0.0491 |
| 16_23007047_G_T | 16 | 23007047 | G/T | 0.1218 | 0.1363 | 0.0927 |
| 16_52538825_C_A | 16 | 52538825 | C/A | 0.1147 | 0.1153 | 0.0917 |
| 16_52599188_C_T | 16 | 52599188 | C/T | 0.1070 | 0.1202 | 0.0431 |
| 16_53809123_C_T | 16 | 53809123 | C/T | -0.0704 | -0.0651 | -0.0957 |
| 16_53861139_C_T | 16 | 53861139 | C/T | -0.0338 | -0.0167 | -0.0782 |
| 16_53861592_G_A | 16 | 53861592 | G/A | -0.0337 | -0.0342 | -0.0303 |
| 16_54682064_G_A | 16 | 54682064 | G/A | 0.0477 | 0.0554 | 0.0304 |
| 16_80648296_A_G | 16 | 80648296 | A/G | 0.0839 | 0.089 | 0.0467 |
| 16_85145977_T_C | 16 | 85145977 | T/C | -0.0211 | -0.0044 | -0.0714 |
| 16_87086492_T_C | 16 | 87086492 | T/C | -0.0469 | -0.0454 | -0.0374 |
| 17_29168077_G_T | 17 | 29168077 | G/T | -0.0568 | -0.0637 | -0.0604 |
| 17_39251123_T_C | 17 | 39251123 | T/C | 0.0799 | 0.0631 | 0.1431 |
| 17_40127060_T_C | 17 | 40127060 | T/C | 0.0174 | -0.0161 | 0.1511 |
| 17_40485239_G_T | 17 | 40485239 | G/T | -0.0571 | -0.0416 | -0.1142 |
| 17_40744470_G_A | 17 | 40744470 | G/A | 0.2017 | 0.1917 | 0.1108 |
| 17_43212339_C_CT | 17 | 43212339 | C/CT | 0.0438 | 0.0352 | 0.0478 |
| 17_44283858_G_A | 17 | 44283858 | G/A | -0.0540 | -0.0532 | -0.0384 |
| 17_53209774_A_C | 17 | 53209774 | A/C | -0.0793 | -0.0933 | -0.0365 |
| 17_77781725_A_G | 17 | 77781725 | A/G | -0.0401 | -0.0378 | -0.0501 |
| 18_11696613_C_T | 18 | 11696613 | C/T | -0.0381 | -0.0281 | -0.094 |
| 18_20634253_C_T | 18 | 20634253 | C/T | -0.0415 | -0.0486 | -0.0284 |

| | | | | | | |
|---|---|---|---|---|---|---|
| 18_24125857_T_C | 18 | 24125857 | T/C | 0.0346 | 0.035 | 0.0231 |
| 18_24337424_C_G | 18 | 24337424 | C/G | 0.0455 | 0.0483 | 0.0282 |
| 18_24518050_AT_A | 18 | 24518050 | AT/A | -0.0599 | -0.083 | 0.006 |
| 18_25407513_C_G | 18 | 25407513 | C/G | 0.0399 | 0.0307 | 0.0648 |
| 18_29981526_G_A | 18 | 29981526 | G/A | -0.1058 | -0.0962 | -0.152 |
| 18_42411803_G_C | 18 | 42411803 | G/C | -0.0877 | -0.1037 | -0.0189 |
| 18_42888797_T_C | 18 | 42888797 | T/C | -0.0542 | -0.0589 | -0.0372 |
| 19_13249921_G_T | 19 | 13249921 | G/T | 0.0956 | 0.0924 | 0.069 |
| 19_17393925_C_A | 19 | 17393925 | C/A | 0.0378 | 0.0036 | 0.1692 |
| 19_18569492_C_T | 19 | 18569492 | C/T | -0.0719 | -0.0778 | -0.0629 |
| 19_19517054_C_CGGGCG | 19 | 19517054 | C/CGGGCG | 0.0437 | 0.0442 | 0.0577 |
| 19_44283031_T_C | 19 | 44283031 | T/C | 0.0619 | 0.0605 | 0.067 |
| 19_46166073_T_C | 19 | 46166073 | T/C | -0.0360 | -0.0447 | -0.0117 |
| 19_55816678_C_T | 19 | 55816678 | C/T | -0.0359 | -0.0381 | -0.0346 |
| 20_5948227_G_A | 20 | 5948227 | G/A | 0.0760 | 0.0762 | 0.0694 |
| 20_11379842_T_C | 20 | 11379842 | T/C | 0.0844 | 0.0998 | 0.0752 |
| 20_41613706_C_G | 20 | 41613706 | C/G | 0.0315 | 0.0266 | 0.0784 |
| 20_52296849_G_A | 20 | 52296849 | G/A | 0.0440 | 0.0539 | 0.0144 |
| 21_16364756_T_G | 21 | 16364756 | T/G | 0.0646 | 0.0742 | 0.0322 |
| 21_16566350_A_G | 21 | 16566350 | A/G | 0.0595 | 0.0678 | 0.0172 |
| 21_16574455_C_A | 21 | 16574455 | C/A | -0.0707 | -0.0808 | -0.0329 |
| 21_47762932_G_A | 21 | 47762932 | G/A | 0.0946 | 0.0852 | 0.089 |
| 22_19766137_C_T | 22 | 19766137 | C/T | -0.0367 | -0.0426 | -0.022 |
| 22_29121087_A_G | 22 | 29121087 | A/G | 0.1839 | 0.2812 | -0.1566 |
| 22_29135543_G_A | 22 | 29135543 | G/A | 0.0654 | 0.0659 | 0.0536 |
| 22_29203724_C_T | 22 | 29203724 | C/T | 0.1405 | 0.1793 | 0.0191 |
| 22_29551872_A_G | 22 | 29551872 | A/G | -0.1716 | -0.1671 | -0.1318 |
| 22_38583315_AAAAG_AAAAGAAAG* | 22 | 38583315 | AAAAG/AAAAGAAAG | -0.0471 | -0.0608 | 0.0079 |
| 22_39343916_T_A | 22 | 39343916 | T/A | 0.0407 | 0.0326 | 0.033 |
| 22_40904707_CT_C | 22 | 40904707 | CT/C | 0.1148 | 0.116 | 0.1203 |
| 22_43433100_C_T | 22 | 43433100 | C/T | -0.0600 | -0.0585 | -0.0515 |
| 22_45319953_G_A | 22 | 45319953 | G/A | -0.0134 | -0.006 | -0.0611 |
| 22_46283297_G_A | 22 | 46283297 | G/A | 0.0736 | 0.0719 | 0.0993 |

*These 7 variants were not available in the UK Biobank

[1]Position build 37

[2]Effect size of the effect allele for overall breast cancer risk, as calcualted by Mavaddat et al., 2019 AJHG

[3]Effect size of the effect allele for ER-positive breast cancer risk, as calcualted by Mavaddat et al., 2019 AJHG

[4]Effect size of the effect allele for ER-negative breast cancer risk, as calcualted by Mavaddat et al., 2019 AJHG

**Supplementary Table 3A:** Mean and SE of the standardized $PRS_{313}$[1] across the countries for overall breast cancer in controls of BCAC dataset, and when adjusted for the first 6 and 10 principal components (PCs).

| Country | Controls | Controls Onco | Controls iCOGS | Cases | Total | Studies | Mean (SE) $PRS_{313}$ adjusted for array[2] | Standard Deviation[3] of the adjusted mean $PRS_{313}$ | Mean of raw $PRS_{313}$ adjusted for array | SE of raw $PRS_{313}$ adjusted for array | Standard Deviation[3] of the adjusted raw mean $PRS_{313}$ | Mean of raw $PRS_{313}$ (not adjusted for array) | Standard Deviation of the raw mean $PRS_{313}$ (not adjusted for array) | Mean (SE) $PRS_{313}$ adjusted for 6 PCs and array | Mean (SE) $PRS_{313}$ adjusted for 10 PCs and array | Mean $PRS_{313}$ Fitted values adjusted for 6 PCs (PRS ~ 6 PCs)[4] |
|---|---|---|---|---|---|---|---|---|---|---|---|---|---|---|---|---|
| Australia | 4049 | 2375 | 1674 | 4662 | 8711 | ABCFS, ABCTB, BCEES, MCCS, KCONFAB/AOCS | -0.005(0.016) | 1.02 | -0.45 | 0.01 | 0.636 | -0.456 | 0.615 | 0.01(0.017) | 0.01(0.017) | -0.005 |
| Belarus | 342 | 249 | 93 | 744 | 1086 | HMBCS | 0.07(0.054) | 1.00 | -0.404 | 0.033 | 0.610 | -0.408 | 0.679 | 0.071(0.057) | 0.058(0.057) | 0.016 |
| Belgium | 1823 | 1268 | 555 | 3398 | 5221 | LMBC | -0.006(0.024) | 1.02 | -0.45 | 0.014 | 0.598 | -0.455 | 0.599 | -0.007(0.024) | 0.002(0.024) | 0.010 |
| Canada | 2277 | 1630 | 647 | 3310 | 5587 | CBCS, MTLGEBCS, OFBCR | 0.018(0.021) | 1.00 | -0.435 | 0.013 | 0.620 | -0.44 | 0.633 | 0.019(0.021) | 0.018(0.021) | 0.013 |
| Denmark | 5241 | 716 | 4525 | 4275 | 9516 | CGPS | -0.013(0.015) | 1.09 | -0.455 | 0.009 | 0.652 | -0.468 | 0.602 | 0.012(0.017) | 0.013(0.018) | -0.031 |
| Finland | 2083 | 422 | 1661 | 2838 | 4921 | HEBCS, KBCP, OBCS | 0.031(0.023) | 1.05 | -0.428 | 0.014 | 0.639 | -0.44 | 0.605 | 0.008(0.044) | 0.042(0.044) | 0.010 |
| France | 1372 | 529 | 843 | 1288 | 2660 | CECILE, EPIC | 0.0003(0.027) | 1.00 | -0.447 | 0.017 | 0.630 | -0.456 | 0.611 | -0.008(0.029) | -0.003(0.029) | 0.008 |
| Germany | 8563 | 4471 | 4092 | 17881 | 26444 | BBCC, BSUCH, EPIC, ESTHER, GC-HBOC, GENICA, GEPARSIXTO, GESBC, HABCS, MARIE, PREFACE, SKKDKFZS, SUCCESSB, SUCCESSC | 0.011(0.011) | 1.02 | -0.44 | 0.007 | 0.648 | -0.447 | 0.608 | 0.004(0.013) | 0.008(0.013) | 0.013 |
| Greece | 607 | 512 | 95 | 1252 | 1859 | CCGP, EPIC, TNBCC | 0.232(0.041) | 1.01 | -0.305 | 0.025 | 0.616 | -0.307 | 0.593 | 0.043(0.046) | 0.043(0.047) | 0.208 |
| Ireland | 719 | 0 | 719 | 793 | 1512 | BIGGS | -0.118(0.038) | 1.02 | -0.519 | 0.023 | 0.617 | -0.534 | 0.620 | -0.015(0.045) | -0.021(0.047) | -0.112 |
| Israel | 724 | 724 | 0 | 1337 | 2061 | BCINIS | 0.047(0.037) | 1.00 | -0.418 | 0.023 | 0.619 | -0.418 | 0.651 | 0.001(0.048) | 0.002(0.048) | 0.062 |
| Italy | 1554 | 1154 | 400 | 1481 | 3035 | EPIC, MBCSG | 0.115(0.025) | 0.99 | -0.376 | 0.016 | 0.631 | -0.38 | 0.606 | -0.007(0.03) | 0.007(0.03) | 0.131 |
| Netherlands | 4407 | 1765 | 2642 | 5016 | 9423 | ABCS, EPIC, ORIGO, RBCS, ABCS-F | 0.021(0.016) | 1.06 | -0.434 | 0.01 | 0.664 | -0.442 | 0.615 | 0.043(0.017) | 0.049(0.018) | -0.019 |
| Norway | 217 | 0 | 217 | 2386 | 2603 | NBCS | 0.077(0.068) | 1.00 | -0.399 | 0.042 | 0.619 | -0.414 | 0.568 | 0.094(0.069) | 0.085(0.07) | -0.027 |
| Poland | 2554 | 2219 | 335 | 2450 | 5004 | PBCS, SZBCS | 0.013(0.02) | 1.01 | -0.439 | 0.012 | 0.606 | -0.441 | 0.621 | 0.025(0.028) | 0.022(0.028) | 0.010 |
| Republic of North Macedonia | 92 | 92 | 0 | 78 | 170 | MABCS | 0.25(0.104) | 1.00 | -0.294 | 0.064 | 0.614 | -0.294 | 0.587 | 0.134(0.105) | 0.138(0.106) | 0.140 |
| Russia | 120 | 120 | 0 | 211 | 331 | HUBCS | 0.18(0.091) | 1.00 | -0.337 | 0.056 | 0.613 | -0.337 | 0.688 | 0.166(0.093) | 0.1(0.094) | 0.044 |
| Spain | 2098 | 1227 | 871 | 3124 | 5222 | BREOGAN, EPIC, HCSC, CNIO-BCS | 0.057(0.022) | 1.01 | -0.412 | 0.013 | 0.595 | -0.418 | 0.603 | -0.006(0.029) | -0.005(0.033) | 0.057 |
| Sweden | 16680 | 9280 | 7400 | 11957 | 28637 | KARBAC, KARMA, MISS, PKARMA, SMC, SASBAC | -0.015(0.008) | 1.03 | -0.456 | 0.005 | 0.646 | -0.462 | 0.615 | 0.005(0.014) | -0.009(0.015) | -0.017 |
| UK | 16854 | 8054 | 8800 | 21331 | 38185 | BBCS, DIETCOMPLYF, EPIC, FHRISK, POSH, PROCAS, SBCS, SEARCH, UKBGS, UKOPS, GLACIER, ICICLE | -0.01(0.009) | 1.17 | -0.453 | 0.005 | 0.649 | -0.46 | 0.611 | 0.019(0.01) | 0.022(0.01) | -0.023 |
| USA | 21696 | 19008 | 2688 | 20448 | 42144 | 2SISTER, AHS, BCFR-NY, BCFR-PA, BCFR-UTAH, CPSII, CTS, MCBCS, MEC, MMHS, MSKCC, NBHS, NC-BCFR, NCBCS, NHS, NHS2, PLCO, SISTER, TNBCC, UBCS, UCIBCS, USRT | 0.029(0.007) | 1.03 | -0.429 | 0.004 | 0.589 | -0.431 | 0.610 | 0.033(0.007) | 0.033(0.007) | 0.013 |
| Total | 94072 | 55815 | 38257 | 110260 | 204332 | | | | | | | | | | | |

[1]PRS was standardized based on the controls of the pooled dataset

[2]Mean $PRS_{313}$ was adjusted for array, and results were fitted without an intercept

[3]Standard deviation was calculated using the SE of the adjusted mean PRS using the formula SD=SE*SQRT(Number_Controls)

[4]Mean $PRS_{313}$ by country using predicted PRS, estimated using linear predictor of PRS vs 6 PCs and the command predict() in R.

**Supplementary Table 3B:** Mean and SE of the standardized $PRS_{313}$[1] across the countries for ER-specific breast cancer in controls of BCAC dataset, and when adjusted for the first 6 or 10 principal components (PCs).

| | | | | | | | ER-positive | | ER-negative | |
|---|---|---|---|---|---|---|---|---|---|---|
| **Country** | **Controls** | **Controls Onco** | **Controls iCOGS** | **Cases** | **Total** | **Studies** | **Mean (SE) $PRS_{313}$ adjusted for array[2]** | **Mean (SE) $PRS_{313}$ adjusted for 6 PCs and array** | **Mean (SE) $PRS_{313}$ adjusted for array[2]** | **Mean (SE) $PRS_{313}$ adjusted for 10 PCs and array** |
| Australia | 4049 | 2375 | 1674 | 4662 | 8711 | ABCFS, ABCTB, BCEES, MCCS, KCONFAB/AOCS | 0.001(0.016) | 0.015(0.017) | -0.029(0.016) | -0.016(0.017) |
| Belarus | 342 | 249 | 93 | 744 | 1086 | HMBCS | 0.062(0.054) | 0.06(0.057) | 0.116(0.054) | 0.104(0.057) |
| Belgium | 1823 | 1268 | 555 | 3398 | 5221 | LMBC | 0.001(0.024) | -0.001(0.024) | -0.033(0.024) | -0.016(0.024) |
| Canada | 2277 | 1630 | 647 | 3310 | 5587 | CBCS, MTLGEBCS, OFBCR | 0.02(0.021) | 0.019(0.021) | 0.003(0.021) | 0.011(0.021) |
| Denmark | 5241 | 716 | 4525 | 4275 | 9516 | CGPS | -0.021(0.015) | 0.008(0.017) | 0.027(0.015) | 0.022(0.018) |
| Finland | 2083 | 422 | 1661 | 2838 | 4921 | HEBCS, KBCP, OBCS | 0.031(0.023) | 0.022(0.044) | 0.021(0.023) | -0.019(0.044) |
| France | 1372 | 529 | 843 | 1288 | 2660 | CECILE, EPIC | 0.008(0.027) | -0.005(0.029) | -0.045(0.027) | -0.026(0.029) |
| Germany | 8563 | 4471 | 4092 | 17881 | 26444 | BBCC, BSUCH, EPIC, ESTHER, GC-HBOC, GENICA, GEPARSIXTO, GESBC, HABCS, MARIE, PREFACE, SKKDKFZS, SUCCESSB, SUCCESSC | 0.017(0.011) | 0.008(0.013) | -0.02(0.011) | -0.012(0.013) |
| Greece | 607 | 512 | 95 | 1252 | 1859 | CCGP, EPIC, TNBCC | 0.257(0.041) | 0.057(0.046) | 0.029(0.041) | -0.017(0.047) |
| Ireland | 719 | 0 | 719 | 793 | 1512 | BIGGS | -0.122(0.038) | -0.018(0.045) | -0.086(0.038) | -0.019(0.047) |
| Israel | 724 | 724 | 0 | 1337 | 2061 | BCINIS | 0.049(0.037) | 0.003(0.048) | -0.05(0.037) | -0.01(0.048) |
| Italy | 1554 | 1154 | 400 | 1481 | 3035 | EPIC, MBCSG | 0.136(0.025) | 0.002(0.03) | -0.029(0.025) | -0.03(0.03) |
| Netherlands | 4407 | 1765 | 2642 | 5016 | 9423 | ABCS, EPIC, ORIGO, RBCS, ABCS-F | 0.02(0.016) | 0.043(0.017) | 0.036(0.016) | 0.051(0.018) |
| Norway | 217 | 0 | 217 | 2386 | 2603 | NBCS | 0.059(0.068) | 0.083(0.069) | 0.118(0.068) | 0.077(0.07) |
| Poland | 2554 | 2219 | 335 | 2450 | 5004 | PBCS, SZBCS | 0.017(0.02) | 0.026(0.028) | 0.002(0.02) | 0.007(0.028) |
| Republic of North Macedonia | 92 | 92 | 0 | 78 | 170 | MABCS | 0.257(0.104) | 0.134(0.105) | 0.168(0.104) | 0.145(0.106) |
| Russia | 120 | 120 | 0 | 211 | 331 | HUBCS | 0.159(0.091) | 0.147(0.093) | 0.232(0.091) | 0.147(0.094) |
| Spain | 2098 | 1227 | 871 | 3124 | 5222 | BREOGAN, EPIC, HCSC, CNIO-BCS | 0.065(0.022) | -0.008(0.029) | 0.006(0.022) | 0.002(0.033) |
| Sweden | 16680 | 9280 | 7400 | 11957 | 28637 | KARBAC, KARMA, MISS, PKARMA, SMC, SASBAC | -0.019(0.008) | 0.007(0.014) | 0.001(0.008) | -0.044(0.015) |
| UK | 16854 | 8054 | 8800 | 21331 | 38185 | BBCS, DIETCOMPLYF, EPIC, FHRISK, POSH, PROCAS, SBCS, SEARCH, UKBGS, UKOPS, GLACIER, ICICLE | -0.011(0.009) | 0.018(0.01) | 0.006(0.009) | 0.025(0.01) |
| USA | 21696 | 19008 | 2688 | 20448 | 42144 | 2SISTER, AHS, BCFR-NY, BCFR-PA, BCFR-UTAH, CPSII, CTS, MCBCS, MEC, MMHS, MSKCC, NBHS, NC-BCFR, NCBCS, NHS, NHS2, PLCO, SISTER, TNBCC, UBCS, UCIBCS, USRT | 0.029(0.007) | 0.033(0.007) | 0.025(0.007) | 0.028(0.007) |

[1]PRS was standardized based on the controls of the pooled dataset

[2]Mean $PRS_{313}$ was adjusted for array, and results were fitted without an intercept

**Supplementary Table 4**: Mean and SE of the standardized $PRS_{306}$[1] and when adjusted for the first 8 or 10 principal components (PCs), and of the standard PRS (as defined by the UK Biobank) across the countries for overall breast cancer, in white females participating in the UK Biobank.

| Country | Mean (SE) $PRS_{306}$ | Mean (SE) $PRS_{306}$ adjusted for 8 PCs | Mean (SE) $PRS_{306}$ adjusted for 10 PCs | Mean (SE) standard PRS |
|---|---|---|---|---|
| Australia | 0.009 (0.044) | 0.051 (0.045) | 0.05 (0.045) | 0.042 (0.044) |
| Austria | -0.048 (0.092) | -0.048 (0.093) | -0.046 (0.093) | -0.046 (0.092) |
| Canada | 0.057 (0.048) | 0.097 (0.049) | 0.096 (0.049) | 0.021 (0.048) |
| Cyprus | 0.274 (0.087) | 0.195 (0.089) | 0.187 (0.089) | 0.404 (0.087) |
| Denmark | 0.159 (0.081) | 0.199 (0.082) | 0.195 (0.082) | 0.056 (0.081) |
| Finland | 0.026 (0.088) | 0.014 (0.092) | 0.012 (0.092) | -0.192 (0.088) |
| France | -0.008 (0.043) | -0.007 (0.045) | 0 (0.045) | 0.053 (0.043) |
| Germany | 0.03 (0.028) | 0.052 (0.03) | 0.053 (0.03) | -0.06 (0.028) |
| Ireland | -0.041 (0.02) | 0.036 (0.023) | 0.031 (0.023) | 0.024 (0.02) |
| Italy | 0.191 (0.054) | 0.098 (0.058) | 0.098 (0.058) | 0.364 (0.054) |
| Malta | 0.193 (0.078) | 0.158 (0.079) | 0.157 (0.079) | 0.319 (0.078) |
| Netherlands | 0.007 (0.057) | 0.044 (0.058) | 0.041 (0.058) | -0.095 (0.057) |
| New Zealand | -0.02 (0.052) | 0.029 (0.053) | 0.027 (0.053) | 0.071 (0.052) |
| Poland | 0.124 (0.05) | 0.108 (0.057) | 0.117 (0.057) | -0.133 (0.05) |
| Portugal | 0.184 (0.075) | 0.105 (0.076) | 0.117 (0.077) | 0.265 (0.075) |
| Russia | 0.267 (0.092) | 0.224 (0.096) | 0.228 (0.096) | -0.082 (0.092) |
| Spain | 0.095 (0.071) | 0.045 (0.072) | 0.059 (0.074) | 0.106 (0.071) |
| Sweden | -0.008 (0.084) | 0.027 (0.085) | 0.022 (0.085) | -0.26 (0.084) |
| Switzerland | 0.013 (0.091) | 0.012 (0.092) | 0.013 (0.092) | 0.127 (0.091) |
| UK, England | 0 (0.002) | 0.047 (0.009) | 0.046 (0.009) | -0.007 (0.002) |
| UK, Northern Ireland | 0.015 (0.026) | 0.081 (0.028) | 0.074 (0.028) | 0.034 (0.026) |
| UK, Scotland | -0.002 (0.007) | 0.059 (0.012) | 0.052 (0.012) | 0.031 (0.007) |
| UK, Wales | -0.022 (0.01) | 0.028 (0.013) | 0.052 (0.014) | 0.027 (0.01) |
| USA | -0.013 (0.035) | 0.013 (0.036) | 0.013 (0.036) | 0.06 (0.035) |

[1]PRS was standardized based on the controls

**Supplementary Table 5:** Mean frequency and standard deviation of each variant in the $PRS_{313}$ across the countries and coefficient of variation.

| SNP name | Mean Frequency | Standard Deviation | Coefficient of variation |
|---|---|---|---|
| 22_29121087_A_G | 0.009 | 0.015 | 1.662 |
| 13_32839990_G_A | 0.017 | 0.012 | 0.692 |
| 5_122705244_C_T | 0.036 | 0.021 | 0.586 |
| 5_1353077_T_C | 0.008 | 0.005 | 0.536 |
| 6_27425644_G_C | 0.067 | 0.032 | 0.473 |
| 22_29135543_G_A | 0.090 | 0.037 | 0.410 |
| 10_22477776_ACC_A | 0.019 | 0.008 | 0.403 |
| 3_29294845_C_T | 0.013 | 0.005 | 0.389 |
| 18_29981526_G_A | 0.047 | 0.018 | 0.380 |
| 3_55970777_A_AT | 0.026 | 0.010 | 0.370 |
| 19_13249921_G_T | 0.041 | 0.014 | 0.345 |
| 21_47762932_G_A | 0.036 | 0.012 | 0.343 |
| 4_38784633_G_T | 0.260 | 0.088 | 0.339 |
| 17_40744470_G_A | 0.011 | 0.004 | 0.331 |
| 1_121287994_A_G | 0.113 | 0.036 | 0.319 |
| 15_46680811_C_A | 0.011 | 0.004 | 0.313 |
| 1_201437832_C_T | 0.055 | 0.017 | 0.311 |
| 22_29203724_C_T | 0.022 | 0.006 | 0.296 |
| 5_345109_T_C | 0.051 | 0.014 | 0.280 |
| 20_5948227_G_A | 0.065 | 0.018 | 0.276 |
| 6_152055978_A_T | 0.063 | 0.017 | 0.274 |
| 1_242034263_A_G | 0.030 | 0.008 | 0.272 |
| 5_56042972_C_T | 0.050 | 0.013 | 0.267 |
| 8_36858483_A_G | 0.199 | 0.053 | 0.265 |
| 1_41389220_T_C | 0.015 | 0.004 | 0.261 |
| 17_44283858_G_A | 0.177 | 0.046 | 0.258 |
| 21_16566350_A_G | 0.086 | 0.022 | 0.258 |
| 10_123349324_A_T | 0.054 | 0.014 | 0.253 |
| 13_32972626_A_T | 0.008 | 0.002 | 0.251 |
| 5_44508264_G_GT | 0.136 | 0.034 | 0.247 |
| 16_23007047_G_T | 0.025 | 0.006 | 0.236 |
| 7_98005235_G_A | 0.159 | 0.037 | 0.235 |
| 10_123340107_A_G | 0.072 | 0.017 | 0.232 |
| 15_67457698_A_G | 0.046 | 0.011 | 0.229 |
| 6_151955914_A_G | 0.075 | 0.017 | 0.228 |
| 3_63887449_T_TTG | 0.120 | 0.027 | 0.225 |
| 9_98362587_T_C | 0.094 | 0.021 | 0.218 |
| 3_87037543_A_G | 0.087 | 0.019 | 0.216 |
| 8_76333056_C_T | 0.091 | 0.019 | 0.212 |
| 11_69331418_C_T | 0.072 | 0.015 | 0.212 |
| 9_110837073_A_G | 0.066 | 0.014 | 0.210 |
| 8_129199566_G_A | 0.184 | 0.038 | 0.208 |
| 6_43227141_G_A | 0.097 | 0.020 | 0.207 |
| 12_28149568_C_T | 0.111 | 0.022 | 0.200 |
| 22_40904707_CT_C | 0.113 | 0.023 | 0.200 |

| | | | |
|---|---|---|---|
| 22_39343916_T_A | 0.251 | 0.049 | 0.197 |
| 2_217955896_GA_G | 0.038 | 0.007 | 0.193 |
| 8_102483100_T_C | 0.096 | 0.018 | 0.189 |
| 1_168171052_CA_C | 0.112 | 0.021 | 0.187 |
| 7_28869017_G_A | 0.108 | 0.020 | 0.187 |
| 2_70172587_G_A | 0.277 | 0.052 | 0.187 |
| 3_46888198_T_C | 0.102 | 0.019 | 0.187 |
| 11_46318032_C_G | 0.063 | 0.012 | 0.185 |
| 17_40127060_T_C | 0.054 | 0.010 | 0.183 |
| 5_79180995_G_GA | 0.171 | 0.031 | 0.180 |
| 3_4742251_A_G | 0.359 | 0.065 | 0.180 |
| 22_43433100_C_T | 0.109 | 0.020 | 0.179 |
| 15_75750383_T_C | 0.271 | 0.048 | 0.179 |
| 22_46283297_G_A | 0.114 | 0.020 | 0.178 |
| 16_53861592_G_A | 0.366 | 0.065 | 0.178 |
| 17_43212339_C_CT | 0.233 | 0.041 | 0.176 |
| 4_143467195_C_T | 0.110 | 0.019 | 0.174 |
| 11_116727936_A_T | 0.203 | 0.035 | 0.173 |
| 2_10138983_T_C | 0.112 | 0.019 | 0.173 |
| 12_120832146_C_T | 0.163 | 0.027 | 0.168 |
| 5_52679539_C_CA | 0.103 | 0.017 | 0.166 |
| 12_115796577_A_G | 0.198 | 0.033 | 0.166 |
| 17_40485239_G_T | 0.082 | 0.014 | 0.165 |
| 12_103097887_C_T | 0.117 | 0.019 | 0.165 |
| 5_44619502_A_G | 0.151 | 0.025 | 0.163 |
| 3_27388664_C_G | 0.280 | 0.045 | 0.160 |
| 2_29179452_G_C | 0.230 | 0.035 | 0.150 |
| 4_106069013_G_T | 0.238 | 0.035 | 0.149 |
| 22_19766137_C_T | 0.396 | 0.058 | 0.147 |
| 18_42411803_G_C | 0.074 | 0.011 | 0.146 |
| 8_120862186_A_G | 0.136 | 0.020 | 0.145 |
| 4_175842495_G_A | 0.113 | 0.016 | 0.145 |
| 3_49709912_C_CT | 0.297 | 0.043 | 0.144 |
| 6_85912194_CAA_C | 0.058 | 0.008 | 0.144 |
| 14_68979835_T_C | 0.250 | 0.036 | 0.144 |
| 7_101552440_G_A | 0.114 | 0.016 | 0.143 |
| 3_141112859_CTT_C | 0.389 | 0.055 | 0.140 |
| 11_108267402_C_CA | 0.434 | 0.060 | 0.138 |
| 10_64299890_A_G | 0.166 | 0.023 | 0.138 |
| 9_110303808_TAA_T | 0.216 | 0.030 | 0.137 |
| 15_100905819_A_C | 0.110 | 0.015 | 0.136 |
| 1_155556971_G_A | 0.224 | 0.030 | 0.136 |
| 18_11696613_C_T | 0.145 | 0.020 | 0.136 |
| 20_52296849_G_A | 0.241 | 0.032 | 0.135 |
| 1_88156923_G_A | 0.146 | 0.020 | 0.134 |
| 5_132407058_C_T | 0.232 | 0.031 | 0.134 |
| 10_5794652_A_G | 0.208 | 0.027 | 0.132 |
| 17_39251123_T_C | 0.064 | 0.008 | 0.132 |
| 2_121089731_T_C | 0.194 | 0.026 | 0.132 |
| 3_172285237_G_A | 0.213 | 0.028 | 0.131 |

| | | | |
|---|---|---|---|
| 9_136146597_C_T | 0.269 | 0.035 | 0.131 |
| 3_30684907_C_T | 0.306 | 0.039 | 0.129 |
| 10_123095209_G_A | 0.340 | 0.044 | 0.129 |
| 1_46670206_TC_T | 0.285 | 0.037 | 0.128 |
| 9_6880263_A_G | 0.288 | 0.037 | 0.127 |
| 1_220671050_C_T | 0.241 | 0.030 | 0.126 |
| 12_83064195_G_GA | 0.101 | 0.013 | 0.124 |
| 12_57146069_T_G | 0.104 | 0.013 | 0.122 |
| 7_91459189_A_ATT | 0.333 | 0.041 | 0.122 |
| 1_118141492_A_C | 0.269 | 0.033 | 0.122 |
| 6_149595505_T_C | 0.208 | 0.025 | 0.121 |
| 5_173358154_G_A | 0.403 | 0.048 | 0.119 |
| 15_50694306_A_G | 0.352 | 0.042 | 0.119 |
| 7_21940960_A_G | 0.348 | 0.041 | 0.119 |
| 5_81512947_TA_T | 0.247 | 0.029 | 0.119 |
| 1_114445880_G_A | 0.166 | 0.020 | 0.117 |
| 1_51467096_CT_C | 0.481 | 0.056 | 0.117 |
| 11_69330983_G_A | 0.122 | 0.014 | 0.116 |
| 12_14413931_G_C | 0.261 | 0.030 | 0.116 |
| 7_25569548_C_T | 0.168 | 0.019 | 0.116 |
| 12_111600134_G_T | 0.375 | 0.043 | 0.116 |
| 5_56045081_T_C | 0.166 | 0.019 | 0.115 |
| 6_151949806_A_C | 0.306 | 0.035 | 0.114 |
| 5_71965007_G_A | 0.248 | 0.028 | 0.114 |
| 12_70798355_A_T | 0.177 | 0.020 | 0.114 |
| 8_106358620_A_T | 0.101 | 0.012 | 0.114 |
| 2_29615233_T_C | 0.274 | 0.031 | 0.114 |
| 10_80886726_A_G | 0.166 | 0.019 | 0.113 |
| 13_73806982_T_C | 0.304 | 0.034 | 0.112 |
| 1_203770448_T_A | 0.267 | 0.030 | 0.111 |
| 5_55662540_C_CT | 0.366 | 0.040 | 0.110 |
| 16_80648296_A_G | 0.237 | 0.026 | 0.110 |
| 11_69328130_A_T | 0.214 | 0.023 | 0.110 |
| 1_7917076_G_A | 0.381 | 0.042 | 0.109 |
| 1_88428199_C_A | 0.247 | 0.027 | 0.109 |
| 1_217220574_G_A | 0.209 | 0.022 | 0.106 |
| 16_87086492_T_C | 0.256 | 0.027 | 0.105 |
| 11_803017_A_G | 0.509 | 0.053 | 0.105 |
| 10_64819996_G_T | 0.198 | 0.021 | 0.104 |
| 6_152432902_C_T | 0.520 | 0.054 | 0.104 |
| 11_65553492_C_A | 0.179 | 0.019 | 0.104 |
| 11_42844441_C_T | 0.322 | 0.033 | 0.104 |
| 16_10706580_G_A | 0.069 | 0.007 | 0.104 |
| 5_104300273_G_T | 0.182 | 0.019 | 0.103 |
| 15_91512267_G_T | 0.139 | 0.014 | 0.103 |
| 19_18569492_C_T | 0.341 | 0.035 | 0.102 |
| 2_218714845_G_A | 0.373 | 0.038 | 0.102 |
| 3_189774456_C_T | 0.216 | 0.022 | 0.102 |
| 12_115108136_T_C | 0.267 | 0.027 | 0.102 |
| 5_77155397_GT_G | 0.349 | 0.035 | 0.101 |

| | | | |
|---|---|---|---|
| 5_90789470_G_A | 0.164 | 0.016 | 0.101 |
| 5_169591460_T_C | 0.342 | 0.034 | 0.100 |
| 7_94113799_T_C | 0.273 | 0.027 | 0.100 |
| 6_16399557_C_T | 0.339 | 0.034 | 0.099 |
| 9_110837176_C_T | 0.164 | 0.016 | 0.097 |
| 14_93070286_C_T | 0.170 | 0.016 | 0.096 |
| 6_169006947_C_G | 0.533 | 0.051 | 0.095 |
| 5_1296255_A_AG | 0.301 | 0.029 | 0.095 |
| 12_28347382_C_T | 0.206 | 0.019 | 0.094 |
| 9_22041998_C_G | 0.140 | 0.013 | 0.093 |
| 4_175847436_C_A | 0.336 | 0.031 | 0.093 |
| 5_44706498_A_G | 0.247 | 0.023 | 0.093 |
| 7_144048902_G_T | 0.227 | 0.021 | 0.092 |
| 1_149906413_T_C | 0.396 | 0.036 | 0.091 |
| 1_208076291_G_A | 0.326 | 0.030 | 0.091 |
| 22_45319953_G_A | 0.418 | 0.038 | 0.091 |
| 2_121159205_G_A | 0.338 | 0.031 | 0.091 |
| 8_23663653_C_A | 0.394 | 0.035 | 0.090 |
| 7_55192256_A_C | 0.536 | 0.048 | 0.089 |
| 8_124563705_T_C | 0.150 | 0.013 | 0.088 |
| 2_172974566_C_G | 0.462 | 0.040 | 0.087 |
| 8_170692_T_C | 0.220 | 0.019 | 0.087 |
| 12_28174817_C_T | 0.239 | 0.021 | 0.086 |
| 17_29168077_G_T | 0.267 | 0.023 | 0.085 |
| 1_10566215_A_G | 0.321 | 0.027 | 0.085 |
| 16_53809123_C_T | 0.429 | 0.036 | 0.084 |
| 9_21964882_CAAAA_C | 0.314 | 0.026 | 0.083 |
| 16_52538825_C_A | 0.263 | 0.022 | 0.083 |
| 5_1279790_C_T | 0.266 | 0.022 | 0.083 |
| 17_53209774_A_C | 0.294 | 0.024 | 0.083 |
| 7_130656911_C_T | 0.362 | 0.030 | 0.083 |
| 16_52599188_C_T | 0.249 | 0.020 | 0.082 |
| 10_71335574_C_T | 0.301 | 0.025 | 0.082 |
| 5_131640536_A_G | 0.535 | 0.043 | 0.080 |
| 12_85004551_C_T | 0.507 | 0.041 | 0.080 |
| 19_17393925_C_A | 0.289 | 0.023 | 0.079 |
| 1_172328767_T_TA | 0.324 | 0.025 | 0.079 |
| 5_56023083_T_G | 0.153 | 0.012 | 0.078 |
| 7_99948655_T_G | 0.216 | 0.017 | 0.078 |
| 6_21923810_T_C | 0.429 | 0.033 | 0.077 |
| 19_19517054_C_CGGGCG | 0.361 | 0.028 | 0.076 |
| 8_76378165_G_T | 0.363 | 0.027 | 0.076 |
| 10_38523626_C_A | 0.361 | 0.027 | 0.075 |
| 1_145604302_C_CT | 0.344 | 0.026 | 0.075 |
| 21_16364756_T_G | 0.167 | 0.012 | 0.074 |
| 6_152023191_G_A | 0.393 | 0.029 | 0.073 |
| 21_16574455_C_A | 0.320 | 0.023 | 0.072 |
| 18_24518050_AT_A | 0.280 | 0.020 | 0.072 |
| 2_19315675_T_A | 0.547 | 0.039 | 0.071 |
| 1_100880328_A_T | 0.407 | 0.029 | 0.071 |

| | | | |
|---|---|---|---|
| 5_44853593_G_C | 0.303 | 0.021 | 0.070 |
| 15_66630569_G_A | 0.626 | 0.043 | 0.069 |
| 1_202184600_C_T | 0.397 | 0.027 | 0.069 |
| 1_121280613_A_G | 0.407 | 0.028 | 0.069 |
| 5_32579616_TCA_T | 0.475 | 0.033 | 0.069 |
| 14_37128564_C_A | 0.210 | 0.014 | 0.068 |
| 22_38583315_AAAAG_AAAAGAAAG | 0.286 | 0.019 | 0.068 |
| 8_128370949_C_G | 0.410 | 0.028 | 0.068 |
| 7_130674481_G_A | 0.299 | 0.020 | 0.068 |
| 2_121058254_A_G | 0.707 | 0.048 | 0.067 |
| 9_119313486_A_G | 0.397 | 0.027 | 0.067 |
| 19_55816678_C_T | 0.356 | 0.023 | 0.064 |
| 19_44283031_T_C | 0.345 | 0.022 | 0.064 |
| 4_89240476_G_A | 0.446 | 0.029 | 0.064 |
| 6_87803819_T_C | 0.271 | 0.017 | 0.064 |
| 7_139943702_CT_C | 0.529 | 0.034 | 0.063 |
| 11_1895708_C_A | 0.398 | 0.025 | 0.063 |
| 16_4106788_C_A | 0.240 | 0.015 | 0.062 |
| 8_128213561_C_CA | 0.409 | 0.025 | 0.062 |
| 11_65572431_G_A | 0.497 | 0.030 | 0.061 |
| 10_22032942_A_G | 0.702 | 0.043 | 0.061 |
| 10_114777670_C_T | 0.462 | 0.028 | 0.060 |
| 2_25129473_A_G | 0.403 | 0.024 | 0.060 |
| 8_128372172_A_G | 0.561 | 0.033 | 0.060 |
| 12_115835836_T_C | 0.414 | 0.025 | 0.059 |
| 5_73234583_T_C | 0.327 | 0.019 | 0.059 |
| 6_82263549_AAT_A | 0.438 | 0.026 | 0.059 |
| 3_71620370_T_G | 0.632 | 0.036 | 0.057 |
| 12_96027759_A_G | 0.296 | 0.017 | 0.057 |
| 1_204502514_T_TTCTGAAACAGGG | 0.806 | 0.046 | 0.057 |
| 11_44368892_G_A | 0.548 | 0.031 | 0.057 |
| 18_24337424_C_G | 0.616 | 0.035 | 0.057 |
| 5_16231194_G_C | 0.560 | 0.032 | 0.057 |
| 3_99403877_G_A | 0.483 | 0.027 | 0.057 |
| 9_110885479_C_T | 0.637 | 0.036 | 0.057 |
| 11_129461016_A_G | 0.608 | 0.034 | 0.056 |
| 2_88358825_G_C | 0.318 | 0.018 | 0.056 |
| 1_217053815_T_G | 0.323 | 0.018 | 0.056 |
| 3_59373745_C_T | 0.422 | 0.024 | 0.056 |
| 5_158244083_C_T | 0.574 | 0.032 | 0.056 |
| 12_293626_A_G | 0.389 | 0.021 | 0.055 |
| 16_85145977_T_C | 0.478 | 0.026 | 0.055 |
| 14_68660428_T_C | 0.821 | 0.045 | 0.054 |
| 4_84370124_TAA_TA | 0.533 | 0.029 | 0.054 |
| 10_13892298_G_A | 0.438 | 0.024 | 0.054 |
| 9_87782211_T_C | 0.516 | 0.028 | 0.054 |
| 7_102481842_T_C | 0.345 | 0.018 | 0.053 |
| 18_24125857_T_C | 0.419 | 0.022 | 0.053 |
| 10_80851257_G_T | 0.625 | 0.033 | 0.053 |
| 14_105213978_T_G | 0.467 | 0.025 | 0.053 |

| | | | |
|---|---|---|---|
| 8_23447496_A_G | 0.656 | 0.035 | 0.053 |
| 5_176134882_T_C | 0.534 | 0.028 | 0.052 |
| 14_37228504_C_T | 0.424 | 0.022 | 0.052 |
| 10_123340431_GC_G | 0.596 | 0.031 | 0.052 |
| 9_36928288_T_C | 0.536 | 0.028 | 0.052 |
| 4_187503758_A_T | 0.446 | 0.023 | 0.052 |
| 4_126752992_A_AAT | 0.519 | 0.026 | 0.051 |
| 17_77781725_A_G | 0.501 | 0.025 | 0.050 |
| 2_217920769_G_T | 0.491 | 0.024 | 0.050 |
| 9_129424719_A_G | 0.454 | 0.023 | 0.050 |
| 6_20537845_CA_C | 0.467 | 0.023 | 0.049 |
| 4_92594859_TTCTTTC_T | 0.445 | 0.022 | 0.049 |
| 6_18783140_G_A | 0.617 | 0.030 | 0.048 |
| 13_73960952_A_G | 0.761 | 0.036 | 0.048 |
| 11_18664241_T_G | 0.710 | 0.034 | 0.047 |
| 18_42888797_T_C | 0.346 | 0.016 | 0.047 |
| 11_103614438_T_G | 0.657 | 0.031 | 0.047 |
| 5_58241712_C_T | 0.580 | 0.027 | 0.046 |
| 8_124571581_G_A | 0.420 | 0.019 | 0.046 |
| 11_122966626_A_G | 0.287 | 0.013 | 0.045 |
| 14_91751788_TC_T | 0.698 | 0.031 | 0.044 |
| 8_117209548_A_G | 0.639 | 0.028 | 0.044 |
| 1_120257110_T_C | 0.531 | 0.023 | 0.044 |
| 8_124739913_T_G | 0.400 | 0.018 | 0.044 |
| 4_151218296_CATATTT_C | 0.655 | 0.029 | 0.044 |
| 8_17787610_CT_C | 0.611 | 0.027 | 0.044 |
| 5_2777029_G_A | 0.412 | 0.018 | 0.044 |
| 18_20634253_C_T | 0.640 | 0.027 | 0.042 |
| 9_110849525_G_T | 0.613 | 0.025 | 0.042 |
| 6_170332621_T_C | 0.605 | 0.025 | 0.041 |
| 16_6963972_C_G | 0.777 | 0.032 | 0.041 |
| 5_44649944_C_T | 0.600 | 0.024 | 0.041 |
| 3_27353716_C_A | 0.531 | 0.021 | 0.040 |
| 2_202204741_T_C | 0.708 | 0.028 | 0.040 |
| 14_91841069_A_G | 0.340 | 0.014 | 0.040 |
| 8_143669254_A_G | 0.355 | 0.014 | 0.039 |
| 19_46166073_T_C | 0.620 | 0.023 | 0.038 |
| 11_433617_T_C | 0.817 | 0.030 | 0.036 |
| 11_111696440_T_C | 0.628 | 0.022 | 0.035 |
| 1_41380440_C_T | 0.651 | 0.022 | 0.034 |
| 16_53861139_C_T | 0.757 | 0.025 | 0.033 |
| 5_122478676_C_A | 0.740 | 0.024 | 0.033 |
| 8_29509616_A_C | 0.671 | 0.022 | 0.033 |
| 6_13713366_G_C | 0.567 | 0.018 | 0.033 |
| 5_55965167_C_T | 0.565 | 0.018 | 0.032 |
| 20_41613706_C_G | 0.805 | 0.025 | 0.031 |
| 16_54682064_G_A | 0.488 | 0.015 | 0.031 |
| 1_18807339_T_C | 0.523 | 0.016 | 0.031 |
| 8_76230943_A_G | 0.830 | 0.024 | 0.029 |
| 6_130341728_C_CT | 0.724 | 0.020 | 0.028 |

| | | | |
|---|---|---|---|
| 10_115128491_T_C | 0.777 | 0.021 | 0.027 |
| 10_95292187_CAA_C | 0.829 | 0.022 | 0.026 |
| 16_4008542_CAAAAA_C | 0.822 | 0.021 | 0.026 |
| 2_174212910_A_G | 0.852 | 0.022 | 0.026 |
| 18_25407513_C_G | 0.703 | 0.018 | 0.025 |
| 6_152022664_CAAAAAAA_C | 0.610 | 0.014 | 0.024 |
| 2_218292158_C_G | 0.728 | 0.017 | 0.023 |
| 2_39699510_C_CT | 0.474 | 0.011 | 0.022 |
| 2_121246568_T_C | 0.893 | 0.019 | 0.022 |
| 13_43501356_A_G | 0.827 | 0.018 | 0.022 |
| 11_129243417_T_G | 0.860 | 0.018 | 0.021 |
| 10_22861490_A_C | 0.935 | 0.019 | 0.020 |
| 1_110198129_CAAA_C | 0.780 | 0.015 | 0.020 |
| 2_192381934_C_T | 0.864 | 0.016 | 0.019 |
| 20_11379842_T_C | 0.950 | 0.017 | 0.018 |
| 12_29140260_G_A | 0.913 | 0.011 | 0.012 |
| 22_29551872_A_G | 0.986 | 0.006 | 0.006 |
| 2_241388857_C_A | 0.978 | 0.006 | 0.006 |

**Supplementary Table 6**: Mean standardized $PRS_{313}$ by country in controls of the pooled BCAC dataset, estimated using an Empirical Bayes approach, and when adjusted for the first 6 principal components (PCs) and array.

| Country | Posterior Mean (no PCs)[1] | Posterior Mean adjusted for 6 PCs[2] | 95% CI Posterior Mean adjusted for 6 PCs[3] | Mean PRS adjusted for 6 PCs[4] |
|---|---|---|---|---|
| Australia | -0.003 | 0.012 | -0.017, 0.041 | 0.010 |
| Belarus | 0.064 | 0.048 | -0.022, 0.119 | 0.071 |
| Belgium | -0.002 | 0.000 | -0.041, 0.041 | -0.007 |
| Canada | 0.020 | 0.020 | -0.017, 0.058 | 0.019 |
| Denmark | -0.012 | 0.014 | -0.012, 0.04 | 0.012 |
| Finland | 0.032 | 0.012 | -0.027, 0.051 | 0.008 |
| France | 0.004 | 0.001 | -0.045, 0.047 | -0.008 |
| Germany | 0.011 | 0.005 | -0.016, 0.025 | 0.004 |
| Greece | 0.199 | 0.038 | -0.023, 0.099 | 0.043 |
| Ireland | -0.092 | 0.002 | -0.056, 0.06 | -0.015 |
| Israel | 0.047 | 0.012 | -0.046, 0.07 | 0.001 |
| Italy | 0.110 | 0.001 | -0.043, 0.045 | -0.007 |
| Netherlands | 0.022 | 0.042 | 0.014, 0.07 | 0.043 |
| Norway | 0.066 | 0.052 | -0.025, 0.129 | 0.094 |
| Poland | 0.015 | 0.026 | -0.01, 0.062 | 0.025 |
| Republic of North Macedonia | 0.129 | 0.049 | -0.037, 0.135 | 0.134 |
| Russia | 0.110 | 0.060 | -0.023, 0.144 | 0.166 |
| Spain | 0.056 | 0.001 | -0.038, 0.039 | -0.006 |
| Sweden | -0.014 | 0.005 | -0.01, 0.02 | 0.005 |
| UK | -0.010 | 0.019 | 0.004, 0.034 | 0.019 |
| USA | 0.029 | 0.033 | 0.02, 0.046 | 0.033 |

[1]Country-specific estimates, means β, using the Empirical Bayes approach, adjusted for array

[2]Country-specific estimates, means β, using the Empirical Bayes approach, adjusted for 6 PCs and array

[3]95% Confidence Interval of posterior mean $PRS_{313}$ adjusted for 6 PCs and array

[4]Mean $PRS_{313}$ adjusted for 6 PCs and array (Supplementary Table 3A)

**Supplementary Table 7A:** Total number and percentage of controls and cases of the BCAC dataset, in each percentile of the standardized $PRS_{313}$ distribution (standardized by the controls). Associations between overall breast cancer risk and $PRS_{313}$ by percentiles.

| Percentiles | Controls | Percentage | Cases | Percentage | OR (95% CI) | Cutoffs (using the controls) |
|---|---|---|---|---|---|---|
| <1% | 941 | 1 | 193 | 0.2 | 0.21 (0.18-0.24) | [-2.3053558, -2.3053558) |
| 1-5% | 3763 | 4 | 1245 | 1.1 | 0.33 (0.31-0.36) | [-2.3053558, -1.6315827) |
| 5-10% | 4704 | 5 | 1969 | 1.8 | 0.42 (0.40-0.45) | [-1.6315827, -1.2814921) |
| 10-20% | 9407 | 10 | 4945 | 4.5 | 0.53 (0.51-0.55) | [-1.2814921, -0.8417227) |
| 20-40% | 18814 | 20 | 13524 | 12.3 | 0.73 (0.70-0.75) | [-0.8417227, -0.2585700) |
| 40-60% | 18814 | 20 | 18651 | 16.9 | REF[1] | [-0.2585700, 0.2491237) |
| 60-80% | 18814 | 20 | 25355 | 23 | 1.36 (1.32-1.40) | [0.2491237, 0.8401592) |
| 80-90% | 9407 | 10 | 17069 | 15.5 | 1.83 (1.77-1.89) | [0.8401592, 1.2827931) |
| 90-95% | 4704 | 5 | 11153 | 10.1 | 2.39 (2.30-2.49) | [**1.2827931**, 1.6581001) |
| 95-99% | 3763 | 4 | 11438 | 10.4 | 3.06 (2.93-3.19) | [**1.6581001**, 2.3365986) |
| ≥99% | 941 | 1 | 4718 | 4.3 | 5.06 (4.70-5.44) | [**2.3365986**, 5.0236339] |

[1] The 40-60% was used as the reference percentile

**Supplementary Table 7B:** Total number and percentage of controls and cases by country of the BCAC dataset, included in the 90th percentile of the distribution when using pooled dataset (Individuals with standardized $PRS_{313} \geq 1.283$)

| Country | Controls | Percentage | Cases | Percentage | Percentage of the diefference | Country | Controls | Cases |
|---|---|---|---|---|---|---|---|---|
| Australia | 439 | 10.842 | 1122 | 24.067 | -0.842 | Australia | 4049 | 4662 |
| Belarus | 52 | **15.205** | 137 | 18.414 | **-5.205** | Belarus | 342 | 744 |
| Belgium | 175 | 9.600 | 770 | 22.660 | 0.400 | Belgium | 1823 | 3398 |
| Canada | 249 | 10.935 | 877 | 26.495 | -0.935 | Canada | 2277 | 3310 |
| Denmark | 487 | 9.292 | 1009 | 23.602 | 0.708 | Denmark | 5241 | 4275 |
| Finland | 230 | 11.042 | 732 | 25.793 | -1.042 | Finland | 2083 | 2838 |
| France | 117 | **8.528** | 315 | 24.457 | **1.472** | France | 1372 | 1288 |
| Germany | 819 | 9.564 | 4549 | 25.440 | 0.436 | Germany | 8563 | 17881 |
| Greece | 83 | **13.674** | 345 | 27.556 | **-3.674** | Greece | 607 | 1252 |
| Ireland | 60 | **8.345** | 157 | 19.798 | **1.655** | Ireland | 719 | 793 |
| Israel | 97 | **13.398** | 348 | 26.028 | **-3.398** | Israel | 724 | 1337 |
| Italy | 183 | 11.776 | 457 | 30.858 | **-1.776** | Italy | 1554 | 1481 |
| Netherlands | 475 | 10.778 | 1394 | 27.791 | -0.778 | Netherlands | 4407 | 5016 |
| Norway | 22 | 10.138 | 589 | 24.686 | -0.138 | Norway | 217 | 2386 |
| Poland | 266 | 10.415 | 622 | 25.388 | -0.415 | Poland | 2554 | 2450 |
| Republic of North Macedonia | 15 | **16.304** | 31 | 39.744 | **-6.304** | Republic of North Macedonia | 92 | 78 |
| Russia | 16 | **13.333** | 56 | 26.540 | **-3.333** | Russia | 120 | 211 |
| Spain | 213 | 10.153 | 839 | 26.857 | -0.153 | Spain | 2098 | 3124 |
| Sweden | 1603 | 9.610 | 2997 | 25.065 | 0.390 | Sweden | 16680 | 11957 |
| UK | 1580 | 9.375 | 5206 | 24.406 | 0.625 | UK | 16854 | 21331 |
| USA | 2227 | 10.265 | 4757 | 23.264 | -0.265 | USA | 21696 | 20448 |

**Supplementary Table 7C**: Total number and percentage of controls and cases by country of the BCAC dataset, included in the 95th percentile of the distribution when using pooled dataset (Individuals with standardized $PRS_{313} \geq 1.658$)

| Country | Controls | Percentage | Cases | Percentage | Percentage of the diefference | Country | Controls | Cases |
|---|---|---|---|---|---|---|---|---|
| Australia | 198 | 4.890 | 687 | 14.736 | 0.110 | Australia | 4049 | 4662 |
| Belarus | 31 | 9.064 | 83 | 11.156 | -4.064 | Belarus | 342 | 744 |
| Belgium | 96 | 5.266 | 471 | 13.861 | -0.266 | Belgium | 1823 | 3398 |
| Canada | 125 | 5.490 | 540 | 16.314 | -0.490 | Canada | 2277 | 3310 |
| Denmark | 243 | 4.637 | 580 | 13.567 | 0.363 | Denmark | 5241 | 4275 |

| | | | | | | | | |
|---|---|---|---|---|---|---|---|---|
| Finland | 114 | 5.473 | 421 | 14.834 | -0.473 | Finland | 2083 | 2838 |
| France | 53 | 3.863 | 188 | 14.596 | 1.137 | France | 1372 | 1288 |
| Germany | 416 | 4.858 | 2686 | 15.022 | 0.142 | Germany | 8563 | 17881 |
| Greece | 38 | 6.260 | 219 | 17.492 | -1.260 | Greece | 607 | 1252 |
| Ireland | 26 | 3.616 | 86 | 10.845 | 1.384 | Ireland | 719 | 793 |
| Israel | 42 | 5.801 | 199 | 14.884 | -0.801 | Israel | 724 | 1337 |
| Italy | 85 | 5.470 | 275 | 18.569 | -0.470 | Italy | 1554 | 1481 |
| Netherlands | 244 | 5.537 | 866 | 17.265 | -0.537 | Netherlands | 4407 | 5016 |
| Norway | 10 | 4.608 | 340 | 14.250 | 0.392 | Norway | 217 | 2386 |
| Poland | 152 | 5.951 | 375 | 15.306 | -0.951 | Poland | 2554 | 2450 |
| Republic of North Macedonia | 8 | 8.696 | 19 | 24.359 | -3.696 | Republic of North Macedonia | 92 | 78 |
| Russia | 7 | 5.833 | 34 | 16.114 | -0.833 | Russia | 120 | 211 |
| Spain | 116 | 5.529 | 515 | 16.485 | -0.529 | Spain | 2098 | 3124 |
| Sweden | 802 | 4.808 | 1783 | 14.912 | 0.192 | Sweden | 16680 | 11957 |
| UK | 787 | 4.670 | 3039 | 14.247 | 0.330 | UK | 16854 | 21331 |
| USA | 1111 | 5.121 | 2750 | 13.449 | -0.121 | USA | 21696 | 20448 |

**Supplementary Table 7D**: Total number and percentage of controls and cases by country of the BCAC dataset, included in the 99th percentile of the distribution when using pooled dataset (Individuals with standardized $PRS_{313}$ ≥ 2.337)

| Country | Controls | Percentage | Cases | Percentage | Percentage of the diefference | Country | Controls | Cases |
|---|---|---|---|---|---|---|---|---|
| Australia | 30 | 0.644 | 201 | 4.311 | 0.356 | Australia | 4049 | 4662 |
| Belarus | 6 | 0.806 | 20 | 2.688 | 0.194 | Belarus | 342 | 744 |
| Belgium | 19 | 0.559 | 148 | 4.356 | 0.441 | Belgium | 1823 | 3398 |
| Canada | 25 | 0.755 | 171 | 5.166 | 0.245 | Canada | 2277 | 3310 |
| Denmark | 39 | 0.912 | 153 | 3.579 | 0.088 | Denmark | 5241 | 4275 |
| Finland | 17 | 0.599 | 122 | 4.299 | 0.401 | Finland | 2083 | 2838 |
| France | 18 | 1.398 | 50 | 3.882 | -0.398 | France | 1372 | 1288 |
| Germany | 92 | 0.515 | 783 | 4.379 | 0.485 | Germany | 8563 | 17881 |
| Greece | 10 | 0.799 | 71 | 5.671 | 0.201 | Greece | 607 | 1252 |
| Ireland | 5 | 0.631 | 27 | 3.405 | 0.369 | Ireland | 719 | 793 |
| Israel | 11 | 0.823 | 56 | 4.188 | 0.177 | Israel | 724 | 1337 |
| Italy | 13 | 0.878 | 96 | 6.482 | 0.122 | Italy | 1554 | 1481 |
| Netherlands | 58 | 1.156 | 270 | 5.383 | -0.156 | Netherlands | 4407 | 5016 |
| Norway | 1 | 0.042 | 91 | 3.814 | 0.958 | Norway | 217 | 2386 |
| Poland | 29 | 1.184 | 102 | 4.163 | -0.184 | Poland | 2554 | 2450 |
| Republic of North Macedonia | 0 | 0.000 | 3 | 3.846 | 1.000 | Republic of North Macedonia | 92 | 78 |
| Russia | 2 | 0.948 | 6 | 2.844 | 0.052 | Russia | 120 | 211 |
| Spain | 19 | 0.608 | 161 | 5.154 | 0.392 | Spain | 2098 | 3124 |
| Sweden | 157 | 1.313 | 529 | 4.424 | -0.313 | Sweden | 16680 | 11957 |
| UK | 151 | 0.708 | 848 | 3.975 | 0.292 | UK | 16854 | 21331 |
| USA | 238 | 1.164 | 809 | 3.956 | -0.164 | USA | 21696 | 20448 |

**Supplementary Table 8:** Total number and percentage of controls and cases in each percentile of the standardized $PRS_{313}$ distribution (standardized by the controls) in Greece, Italy and Ireland of the BCAC dataset. Associations between overall breast cancer risk and $PRS_{313}$ by percentiles.

**Greece**

| Percentiles | Controls | Percentage | Cases | Percentage | OR (95% CI) | Cut-offs values |
|---|---|---|---|---|---|---|
| <1% | 7 | 1.2 | 8 | 0.6 | 0.63 (0.22-1.83) | [min, -1.88176607) |
| 1-5% | 24 | 4 | 29 | 2.3 | 0.66 (0.37-1.20) | [-1.88176607, -1.24389310) |
| 5-10% | 30 | 4.9 | 20 | 1.6 | 0.37 (0.20-0.67) | [-1.24389310, -1.01592260) |
| 10-20% | 61 | 10 | 61 | 4.9 | 0.55 (0.36-0.84) | [-1.01592260, -0.61338082) |
| 20-40% | 121 | 19.9 | 172 | 13.7 | 0.78 (0.57-1.08) | [-0.61338082, -0.03013492) |
| 40-60% | 121 | 19.9 | 220 | 17.6 | REF[1] | [-0.03013492, 0.47473869) |
| 60-80% | 121 | 19.9 | 301 | 24 | 1.37 (1.01-1.86) | [0.47473869, 1.07401938) |
| 80-90% | 61 | 10 | 152 | 12.1 | 1.37 (0.95-1.99) | [1.07401938, 1.44121042) |
| 90-95% | 30 | 4.9 | 97 | 7.7 | 1.78 (1.13-2.87) | [1.44121042, 1.74175509) |
| 95-99% | 24 | 4 | 143 | 11.4 | 3.28 (2.05-5.43) | [1.74175509, 2.54277555) |
| ≥99% | 7 | 1.2 | 49 | 3.9 | 3.85 (1.80-9.55) | [2.54277555, max] |

**Ireland**

| Percentiles | Controls | Percentage | Cases | Percentage | OR (95% CI) | Cut-offs values |
|---|---|---|---|---|---|---|
| <1% | 8 | 1.1 | 0 | 0 | na | [min, -2.78568527) |
| 1-5% | 28 | 3.9 | 10 | 1.3 | 0.43 (0.19-0.90) | [-2.78568527, -1.85698670) |
| 5-10% | 36 | 5 | 8 | 1 | 0.27 (0.11-0.57) | [-1.85698670, -1.41687903) |
| 10-20% | 72 | 10 | 37 | 4.7 | 0.62 (0.39-0.99) | [-1.41687903, -0.88761545) |
| 20-40% | 144 | 20 | 95 | 12 | 0.80 (0.56-1.14) | [-0.88761545, -0.35313128) |
| 40-60% | 143 | 19.9 | 118 | 14.9 | REF[1] | [-0.35313128, 0.09498293) |
| 60-80% | 144 | 20 | 201 | 25.3 | 1.69 (1.22-2.34) | [0.09498293, 0.67706851) |
| 80-90% | 72 | 10 | 146 | 18.4 | 2.46 (1.70-3.58) | [0.67706851, 1.17758635) |
| 90-95% | 36 | 5 | 63 | 7.9 | 2.12 (1.32-3.44) | [1.17758635, 1.50563968) |
| 95-99% | 28 | 3.9 | 79 | 10 | 3.42 (2.11-5.68) | [1.50563968, 2.15571744) |
| ≥99% | 8 | 1.1 | 36 | 4.5 | 5.45 (2.56-13.04) | [2.15571744, 2.77218282] |

**Italy**

| Percentiles | Controls | Percentage | Cases | Percentage | OR (95% CI) | Cut-offs values |
|---|---|---|---|---|---|---|
| <1% | 16 | 1 | 3 | 0.2 | 0.24 (0.06-0.74) | [min., -2.1102335) |
| 1-5% | 62 | 4 | 10 | 0.7 | 0.21 (0.10-0.40) | [-2.1102335, -1.5322247) |
| 5-10% | 78 | 5 | 33 | 2.2 | 0.55 (0.35-0.85) | [-1.5322247, -1.1617753) |
| 10-20% | 155 | 10 | 63 | 4.3 | 0.53 (0.37-0.74) | [-1.1617753, -0.7342892) |
| 20-40% | 311 | 20 | 172 | 11.6 | 0.72 (0.56-0.92) | [-0.7342892, -0.1293942) |
| 40-60% | 310 | 19.9 | 239 | 16.1 | REF[1] | [-0.1293942, 0.3798952) |
| 60-80% | 311 | 20 | 320 | 21.6 | 1.33 (1.06-1.68) | [0.3798952, 0.9339705) |
| 80-90% | 155 | 10 | 228 | 15.4 | 1.91 (1.47-2.49) | [0.9339705, 1.3848687) |
| 90-95% | 78 | 5 | 154 | 10.4 | 2.56 (1.86-3.54) | [1.3848687, 1.7086773) |
| 95-99% | 62 | 4 | 156 | 10.5 | 3.26 (2.34-4.61) | [1.7086773, 2.2943850) |
| ≥99% | 16 | 1 | 103 | 7 | 8.35 (4.94-15.03) | [2.2943850, 5.0236339] |

[1] The 40-60% was used as the reference percentile

**Supplementary Table 9:** Associations between $PRS_{313}$ and breast cancer risk by country, when using the pooled BCAC dataset.

| Country | OR (95% CI)[1] | OR (95% CI)[2] adjusted by 10 PCs |
|---|---|---|
| All | 1.80 (1.78-1.82) | 1.80 (1.78-1.82) |
| Australia | 1.83 (1.74-1.92) | 1.83 (1.75-1.93) |
| Belarus | 1.37 (1.20-1.58) | 1.39 (1.21-1.61) |
| Belgium[3] | 1.80 (1.68-1.92) | 1.85 (1.70-2.02) / 1.79 (1.68-1.92) |
| Canada | 1.77 (1.67-1.88) | 1.77 (1.66-1.88) |
| Denmark | 1.80 (1.72-1.88) | 1.80 (1.72-1.88) |
| Finland | 1.86 (1.75-1.98) | 1.86 (1.75-1.99) |
| France | 1.83 (1.68-1.99) | 1.85 (1.70-2.01) |
| Germany | 1.83 (1.78-1.88) | 1.83 (1.78-1.88) |
| Greece | 1.66 (1.50-1.85) | 1.65 (1.49-1.84) |
| Ireland | 1.89 (1.69-2.12) | 1.93 (1.72-2.16) |
| Israel | 1.73 (1.57-1.90) | 1.73 (1.57-1.90) |
| Italy | 1.91 (1.76-2.06) | 1.91 (1.76-2.07) |
| Netherlands | 1.95 (1.86-2.04) | 1.95 (1.86-2.04) |
| Norway[4] | 1.75 (1.51-2.03) | 1.81 (1.54-2.13) |
| Poland | 1.79 (1.69-1.90) | 1.79 (1.69-1.91) |
| Republic of North Macedonia | 1.75 (1.29-2.44) | 1.87 (1.34-2.67) |
| Russia | 1.59 (1.27-2.01) | 1.59 (1.26-2.03) |
| Spain | 1.88 (1.77-2.00) | 1.88 (1.77-2.00) |
| Sweden | 1.86 (1.81-1.91) | 1.86 (1.81-1.91) |
| UK | 1.85 (1.81-1.89) | 1.85 (1.81-1.89) |
| USA | 1.67 (1.63-1.70) | 1.66 (1.63-1.70) |

[1] Adjusted for array and study when appropriate.

[2] Adjusted for the first 10 PCs of iCOGS and 10 PCs of OncoArray, and array and study when appropriate.

[3] For Belgium, in the second column, the first value was derived when the model was also adjusted for the LMBC PC, and the second when was not.

[4] Not adjusted for array, since OncoArray did not include controls

**Supplementary Table 10:** Associations between $PRS_{313}$ and breast cancer risk by country, when excluding samples included in the training set of BCAC (as used from Mavaddat et al., 2019 AJHG (1)).

| Country | Controls | Cases | OR (95% CI)[1] | OR (95% CI)[2] adjusted for 10 PCs | Mean (SE) PRS in Controls |
|---|---|---|---|---|---|
| All | 28098 | 26618 | 1.64 (1.61-1.67) | 1.64 (1.61-1.67) | 0.047 |
| Australia* | 383 | 411 | 1.46 (1.25-1.71) | 1.51 (1.29-1.77) | 0.081(0.05) |
| Belgium | 87 | 156 | 1.56 (1.19-2.07) | 1.72 (1.28-2.34) | 0.112(0.11) |
| Canada | 432 | 168 | 1.61 (1.34-1.95) | 1.64 (1.36-1.99) | 0.2(0.05) |
| Denmark | 142 | 230 | 1.73 (1.37-2.21) | 1.80 (1.42-2.32) | -0.002(0.08) |
| France | 370 | 378 | 1.59 (1.38-1.84) | 1.60 (1.39-1.86) | 0.097(0.05) |
| Germany | 928 | 4118 | 1.59 (1.44-1.75) | 1.58 (1.48-1.68) | 0.041(0.03) |
| Greece | 246 | 306 | 1.61 (1.35-1.94) | 1.63 (1.36-1.97) | 0.3(0.06) |
| Israel | 145 | 263 | 1.63 (1.33-2.01) | 1.64 (1.33-2.05) | 0.115(0.08) |
| Italy* | 788 | 744 | 1.78 (1.60-1.98) | 1.78 (1.60-1.98) | 0.07(0.04) |
| Netherlands* | 808 | 782 | 1.54 (1.39-1.71) | 1.54 (1.39-1.71) | 0.047(0.04) |
| Poland | 331 | 217 | 1.57 (1.33-1.87) | 1.57 (1.32-1.87) | -0.001(0.06) |
| Spain | 647 | 812 | 1.61 (1.44-1.79) | 1.63 (1.47-1.83) | 0.061(0.04) |
| Sweden | 5052 | 1819 | 1.68 (1.59-1.79) | 1.68 (1.58-1.78) | 0.0003(0.01) |
| UK | 4503 | 5082 | 1.60 (1.51-1.69) | 1.61 (1.52-1.70) | 0.069(0.02) |
| USA | 13236 | 11132 | 1.64 (1.59-1.68) | 1.64 (1.59-1.69) | 0.047(0.01) |

$PRS_{313}$ was standardized based on the mean and SD of all controls in the dataset

[1] Adjusted for array and study when appropriate.

[2] Adjusted for the first 10 PCs of iCOGS and 10 PCs of OncoArray, and array and study when appropriate.

*Only few individuals genotyped using iCOGS, so not adjusted for array (and icogs' PCs). For Italy, also not adjusted for study.
